# Adaptive Optics Fluorescence Lifetime Imaging Ophthalmoscopy for Single-Cell–Resolved In Vivo Metabolic and Structural Imaging of the Human Retinal Pigment Epithelium

**DOI:** 10.64898/2026.02.07.26345814

**Authors:** Ruixue Liu, Xiaolin Wang, Giulia Corradetti, Ceren Soylu, Deborah A. Ferrington, Srinivas R Sadda, Yuhua Zhang

**Affiliations:** Doheny Eye Institute, Pasadena, California, United States; The Department of Ophthalmology, University of California, Los Angeles, California, United States

## Abstract

Fluorescence lifetime imaging ophthalmoscopy permits in vivo assessment of retinal metabolism but has remained limited by insufficient cellular resolution in the human eye. Here we present adaptive optics–enhanced fluorescence lifetime imaging ophthalmoscopy (AOFLIO), a method for single-cell–resolved, in vivo structural and metabolic imaging of the human retinal pigment epithelium (RPE). Through real-time correction of ocular wavefront aberrations, precisely synchronized adaptive optics reflectance and lifetime image acquisition via a phase-locked loop–based timing architecture, and sub-pixel photon registration that localizes individual autofluorescence photons with high spatial precision, AOFLIO directly resolves the RPE cell mosaic and measures autofluorescence decay using the same photons, enabling direct structural–functional correlation at the single-cell level. We demonstrate single-cell RPE lifetime mapping in healthy subjects and reveal altered metabolic signatures and fine characterization of RPE metabolic in age-related macular degeneration. AOFLIO establishes a platform for cellular-scale metabolic imaging in the living human eye.

## 1. Introduction

The optical accessibility of the retina, together with the characteristic absorbance and fluorescence properties of many endogenous fluorophores,^1,2^ has motivated the development of quantitative imaging approaches to interrogate retinal physiology and pathology.^3–11^ These intrinsic fluorophores arise from metabolic activity and intracellular processing within the neural retina and retinal pigment epithelium (RPE), ranging from vitamin A derivatives in lipofuscin (LF) to flavin adenine dinucleotide (FAD) in mitochondria,^12–21^ providing a biological basis for noninvasive assessment of cellular function beyond structural imaging.^11^

Fluorescence lifetime is a measure of the duration that a fluorophore remains in the excited state before returning to the ground state by emitting a photon.^22^ It is an intrinsic property unique to each fluorescent molecule and its environmental state.^22^ Thus, fluorescence lifetime imaging offers a quantitative and bioenvironmentally sensitive contrast mechanism that surpasses intensity-based measurements.^22,23^ Fluorescence lifetime imaging ophthalmoscopy (FLIO) has been demonstrated to measure the fundus autofluorescence lifetime in the living human eye, providing a functional readout of metabolic state in the living human eye.^7,10,24^ Applications of FLIO across a range of retinal and neurodegenerative conditions, including age-related macular degeneration (AMD),^25–27^ diabetic retinopathy,^28^ central serous chorioretinopathy,^29^ retinitis pigmentosa,^30^ choroideremia,^31^ Stargardt Disease,^32^ Macular Telangiectasia 2,^33,34^ and Alzheimer’s disease,^35^ have demonstrated its translational potential. However, current FLIO implementations are fundamentally limited by ocular aberrations and coarse spatial resolution, resulting in tissue-averaged measurements that obscure cellular heterogeneity.

Adaptive optics (AO) corrects ocular aberrations and enables diffraction-limited retinal imaging,^36,37^ but integrating AO with fluorescence lifetime imaging in the living human eye faces substantial technical challenges, including synchronization of raster scanning with time-correlated single-photon counting, accurate photon localization in the presence of nonlinear scanning trajectory and involuntary eye motion.^38–41^ Consequently, although AO-enhanced FLIO has been demonstrated, it has not achieved robust single-cell–resolved lifetime imaging of the human RPE, particularly in the metabolically informative 500–560 nm spectral range associated with FAD.^40^

This limitation is especially consequential for studying RPE, a central regulator of chorioretinal health through essential functions including retinoid recycling, phagocytosis of shed photoreceptor outer segments, oxidative stress regulation, and metabolic exchange with the choroid.^42–46^ Dysregulation of these processes is a key contributor to a wide range of retinal diseases and often precedes overt structural degeneration. RPE is enriched in intrinsic fluorophores, including lipofuscin components, FAD, and other metabolic cofactors, whose fluorescence lifetimes report molecular composition, redox state, and microenvironment.^2,4,11,47,48^ As these fluorophores evolve with aging and disease,^49^ their lifetime signatures provide quantitative biomarkers of RPE metabolic function.

Here, we present an AOFLIO framework that overcomes these limitations and enables robust, single-cell–resolved fluorescence lifetime imaging of the human RPE in vivo. The system integrates a phase-locked loop (PLL) based timing architecture with non-interpolative, subpixel photon localization and high-precision eye-motion correction, enabling accurate spatiotemporal assignment of individual autofluorescence photons. By reconstructing RPE cellular structure directly from the same photons used for lifetime analysis, AOFLIO enables direct structural–functional correlation at the cellular scale, establishing a new platform for in vivo interrogation retinal metabolism.

## 2. Results

### 2.1 AOFLIO: a precisely synchronized multimodal imaging system

AOFLIO integrates an adaptive optics scanning laser ophthalmoscope (AOSLO)^50^ with a fluorescence lifetime imaging pathway based on time-correlated single-photon counting (TCSPC). This configuration ensures precise spatial registration of individual autofluorescence photons and accurate measurement of autofluorescence lifetime (**Fig. 1**).

**Fig. 1.**
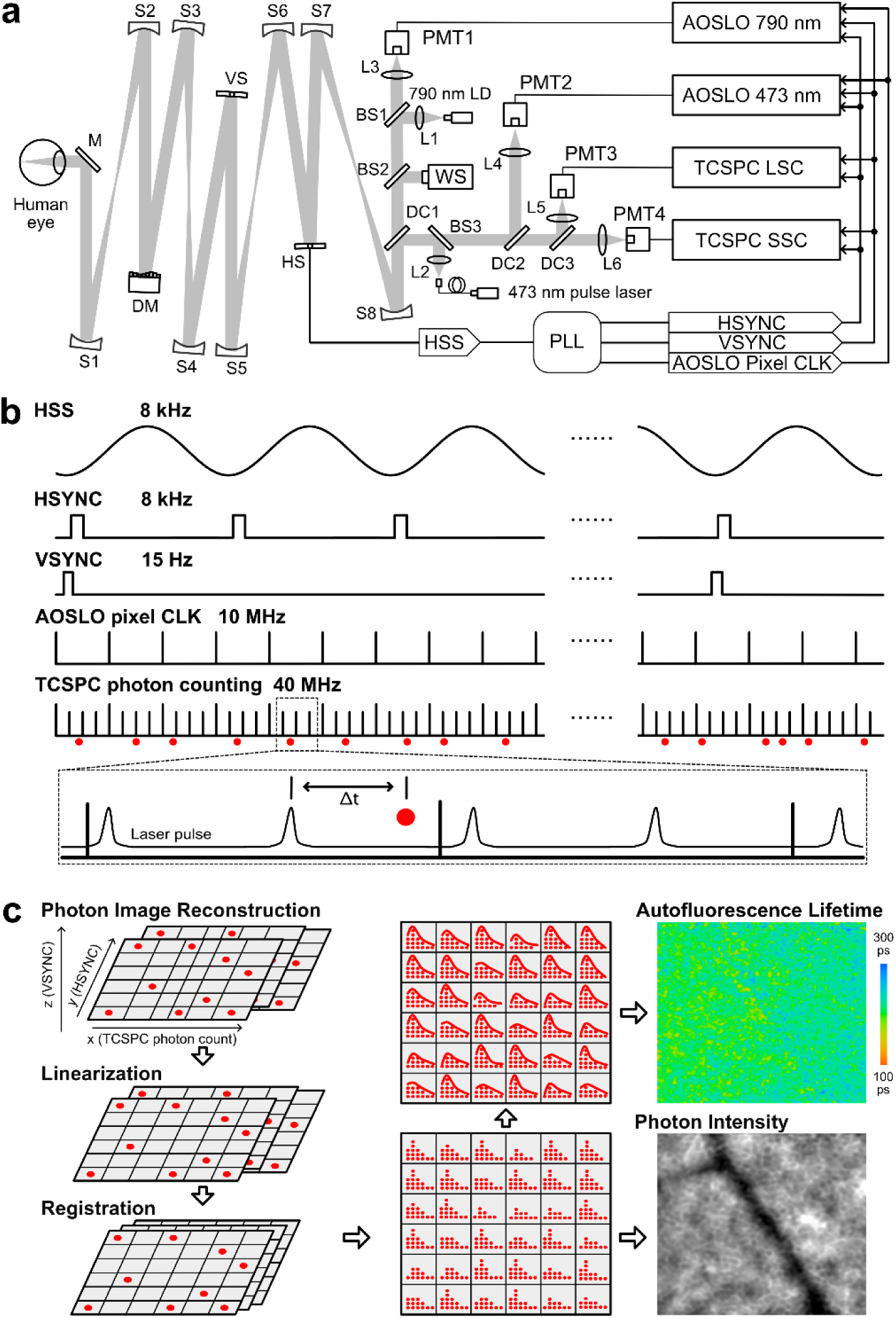
Adaptive optics fluorescence lifetime imaging ophthalmoscopy (AOFLIO) with synchronized multi-channel acquisition of retinal reflectance and autofluorescence and spatiotemporal photon reconstruction. **a,** AOFLIO optical and electronic schematic system showing four-channel synchronized acquisition. AOSLO reflectance images at 790 nm and 473 nm, together with autofluorescence lifetime imaging channels in the long spectral channel (LSC; 560–720 nm) and short spectral channel (SSC; 498–560 nm), are acquired using the same line (HSYNC) and frame synchronization (VSYNC). These signals are derived from the horizontal scan signal (HSS) via a phase-locked loop (PLL), ensuring precise tracking of the scanning trajectory. **b,** Timing diagram illustrating system clocks and photon counting. AOSLO imaging operates at a 10 MHz pixel rate, while AOFLIO time-correlated single-photon counting (TCSPC) operates at 40 MHz, enabling hardware-level pixel subdivision. Detected photons are time-tagged relative to the excitation laser pulse (micro-time, Δt), the start of image acquisition (macro-time), the raster scanning HSYNC and VSYNC. **c,** Spatiotemporal reconstruction of detected autofluorescence photons into two-dimensional images using synchronized line and frame signals in combination with autofluorescence photon macro-time. Photon positions are deterministically assigned to correct for resonant scan induced distortion and subsequently registered using eye-motion estimates derived from simultaneously acquired AOSLO (λ = 790 nm) reflectance images. Photon counts yield autofluorescence intensity images, whereas photon time tags are used to reconstruct fluorescence decay and produce lifetime maps. Details of the non-interpolative pixel subdivision and photon reassignment procedure are provided in **Extended Data Fig. 1. M**, mirror; S1-S8, spherical mirrors; BS1-BS3, beam splitters; L1-L6, lenses; DC1-DC3, dichroic mirrors; SLD, superluminant diode; PMT1-PMT4, photon multipliers.

AOSLO employed a near-infrared (NIR) superluminescent diode (SLD; λ = 790 nm; HS-790, Superlum Diodes Ltd., Cork, Ireland) for reflectance imaging and wavefront sensing. Light was coupled into the scanning system via beam splitter BS1. The scanning optics consisted of a series of spherical mirrors (S1–S8), a resonant scanner for horizontal scanning (HS), a galvanometric scanner for vertical scanning (VS), and a deformable mirror (DM97-15, Bertin Alpao, Montbonnot, France), relaying the beam onto the retina. Retinal reflectance retraced the incoming optical path back to BS1, where 90% of the light was transmitted to the imaging channel and detected by a photomultiplier tube (PMT; H7422-50, Hamamatsu Photonics, Japan). Images were recorded using a four-channel frame grabber (SOL 6M 4AE, Matrox Electronic Systems Ltd., Quebec, Canada). Adaptive optics correction was achieved using a custom wavefront sensor and deformable mirror operating in closed loop at 20–100 Hz with custom control software.^51–53^

AOFLIO was implemented by introducing a picosecond pulsed diode laser (λ = 473 nm) (BDS-SM, Becker & Hickl GmbH, Germany) as the retinal autofluorescence excitation source. Excitation light was coupled into the scanning optical system via beam splitter BS3 and a dichroic mirror (DC1), following the same scanning path as the AOSLO beam. Retinal reflectance and autofluorescence retraced the optical path back to DC1 and were directed into the AOFLIO detection pathway. At beam splitter BS3, 90% of the returned light was transmitted to the imaging channels. Reflectance at 473 nm was detected by a second PMT (H7422-40, Hamamatsu Photonics) and acquired by the AOSLO frame grabber.

Autofluorescence was detected in a long spectral channel (LSC, 560–720 nm) and a short spectral channel (SSC, 500–560 nm) using single-photon counting detectors (HPM-100-40, Becker & Hickl GmbH) connected to a dual-channel TCSPC module (SPC-180NX, Becker & Hickl GmbH).

A custom PLL based timing system was developed to track resonant scanner motion with phase jitters below 50 ns, generating the line and frame synchronization signals as well as AOSLO pixel clock.^54^ This timing architecture synchronized AOSLO reflectance imaging at both 790 nm and 473 nm with AOFLIO TCSPC in both the LSC and SCC. AOSLO reflectance images were acquired with a 10 MHz pixel clock, producing 512 × 512 pixels frames (0.67 μm/pixel) at 15 Hz over a 1.2° × 1.2° field of view. AOFLIO acquisition was synchronized to the same scan lines and frames but operated at a fourfold higher pixel clock (40 MHz), enabling subpixel subdivision of AOSLO pixels. Autofluorescence excitation was delivered at 80 MHz.

Because the AOSLO frame grabber and TCSPC module operated with independent clocks, the AOSLO pixel clock was carefully adjusted to match the TCSPC timing, with a maximum relative timing error of one 80 MHz pulse period. Although the AOSLO clock could be used to drive the TCSPC, the TCSPC internal clock was derived directly from the laser pulse train and provided superior timing accuracy and maximum photon detection probability. This acquisition scheme enabled localization of individual autofluorescence photons with a spatial precision of approximately 0.17 µm on the retina, exceeding the native spatial sampling of the AOSLO structural images. Each detected photon was tagged with its arrival time relative to the excitation pulse (micro-time), the corresponding line and frame synchronization signals, and the elapsed time from acquisition start (macro-time), thereby precisely defining both fluorescence lifetime and spatial origin.^23^

Sinusoidal distortion induced by the resonant scanner was linearized using a non-interpolative linearization approach (**Methods 4.3**) that reassigned individual autofluorescence photons to their corrected retinal coordinates without fractional redistribution, preserving photon statistics and lifetime information. Retinal motion was estimated from simultaneously acquired AOSLO (λ = 790 nm) images using a high-precision substrip-based registration algorithm.^55^ The calculated motion traces were applied to AOSLO reflectance at 473 nm and AOFLIO data, registering autofluorescence photons to their corrected spatial origins. At each registered pixel location, autofluorescence photon counts were averaged to generate autofluorescence intensity images, while the distribution (histogram) of photon arrival times defined the corresponding autofluorescence decay.

### 2.2 AOFLIO Produced Cellular-Resolution Imaging of RPE Structure and Metabolic Function in the Living Human Eye

AOFLIO imaged 34 eyes of 34 subjects with normal chorioretinal health (age: 50.3LJ±LJ16.6LJyears; range: 23–82 years; **Supplementary Table 1**). Each subject was imaged at 5 - 6 non-overlapping sites within 0 - 10° nasal eccentricity. AOFLIO directly visualized individual RPE cells using the autofluorescence photons acquired for measuring fluorescence lifetime (**Fig. 2**). The clear depiction of cell structures indicates that the autofluorescence photons originated predominantly from the RPE cells, thereby facilitating a direct structural-functional correlation at the single-cell level.

**Fig. 2.**
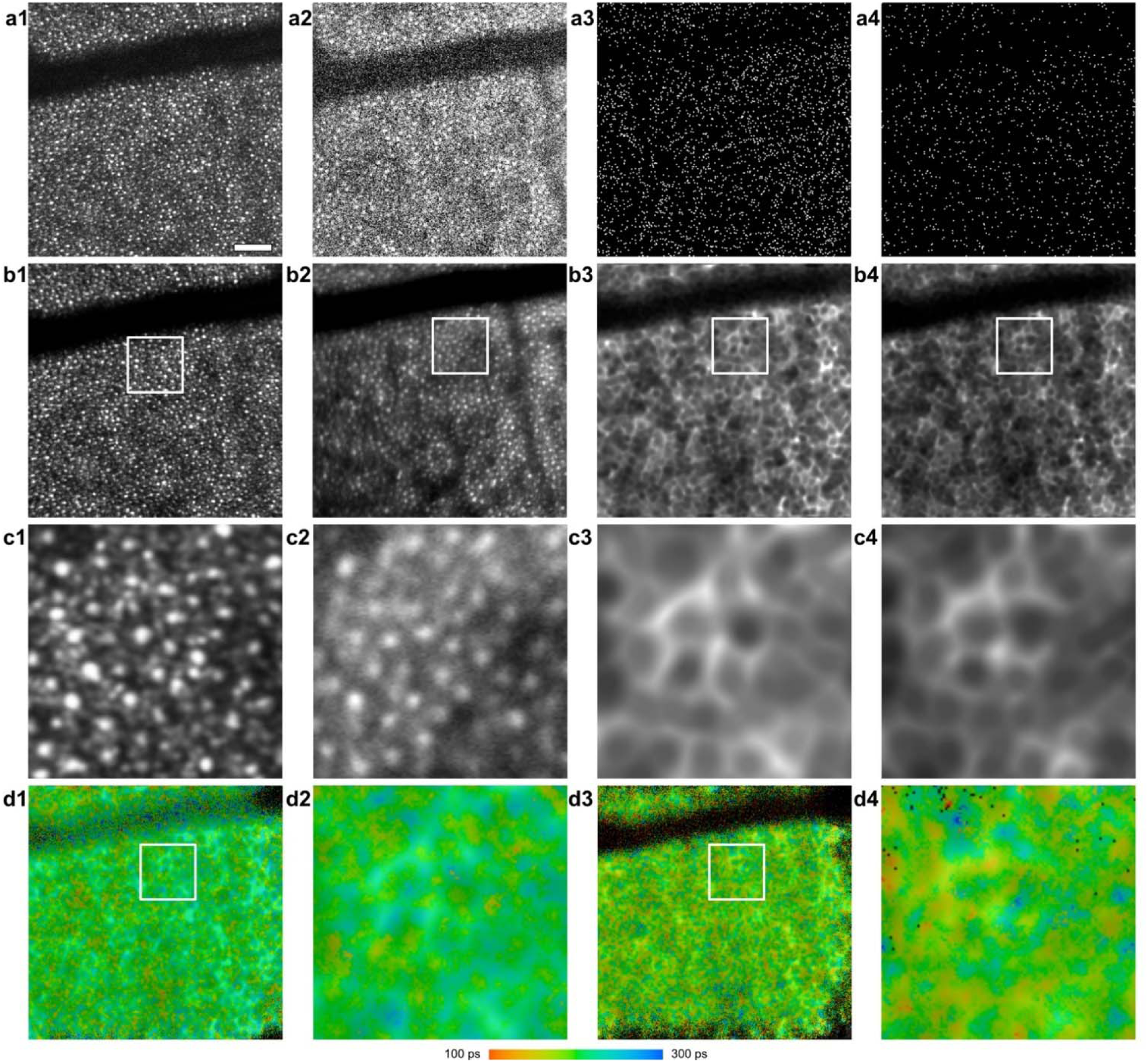
Adaptive optics fluorescence lifetime imaging ophthalmoscopy (AOFLIO) of photoreceptor and retinal pigment epithelium (RPE) cells. **a1**–**a4,** Single-frame images acquired simultaneously: AOSLO reflectance at 790 nm, AOSLO reflectance at 473 nm, AOFLIO LSC, and AOFLIO SSC, respectively. **b1**–**b4,** Corresponding spatially registered images in the same order. **c1**–**c4,** Magnified views (0.3°L×L0.3°) of the corresponding selected regions shown in panels b1- b4. **d1**, RPE autofluorescence lifetime measured in the long spectral channel. **d2**, magnified view of the selected region indicated in panel d1. **d3**, RPE autofluorescence lifetime measured in the short spectral channel. **d2**, magnified view of the selected region indicated in panel d4. These images were acquired from a healthy subject in their third decade of life at 8° eccentricity nasally. Scale bar, 50 µm.

AOFLIO enabled subcellular-scale imaging that revealed spatial heterogeneity in fluorescence lifetime signatures within individual RPE cells (**Fig. 3**). Using a robust coefficient of variation (rCV, IQR/median; see Methods), intra-cell dispersion assessed within 0.3°LJ×LJ0.3° regions was 3.47LJ±LJ1.68%, 6.60LJ±LJ3.85%, and 12.39LJ±LJ6.56% for τ_m12_, τ_1_, and τ_2_ in the LSC, respectively, and 3.62 ± 2.68 % and 11.32 ± 11.32 % for α_1_ and α_2_. Corresponding values in the SSC were 5.32 ± 2.49 %, 10.56 ± 6.26 %, and 21.47 ± 9.79 % for τ_m12_, τ_1_, and τ_2_, and 6.78 ± 4.88 % and 29.15 ± 14.23 % for α_1_ and α_2_.

**Fig. 3.**
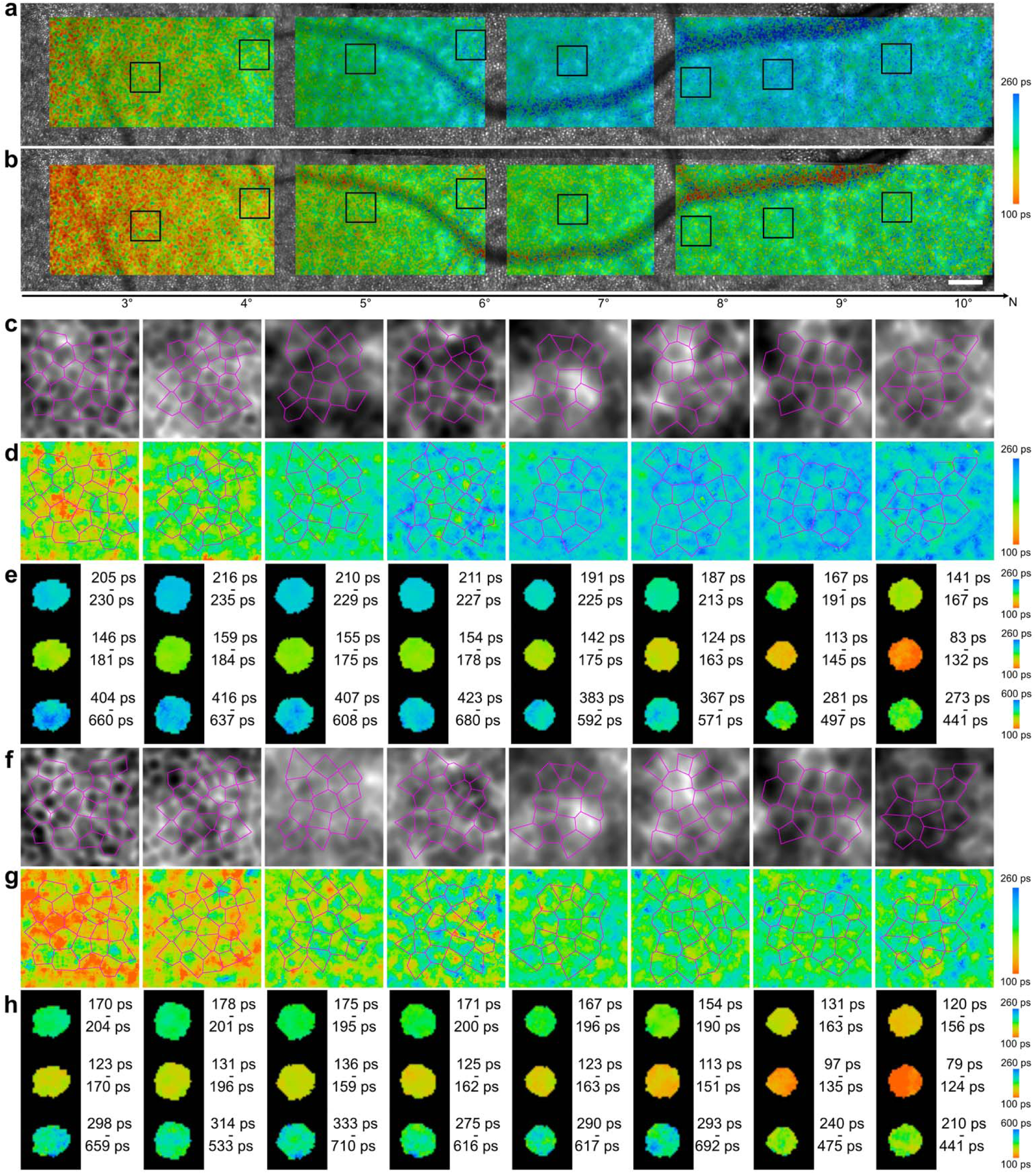
Adaptive optics fluorescence lifetime imaging ophthalmoscopy (AOFLIO) of subcellular-level retinal pigment epithelium (RPE) autofluorescence lifetime in a healthy subject. **a & b**, Autofluorescence lifetime in LSC and SSC overlaid on adaptive optics reflectance image (N: Nasal; scale bar, 0.3°). **c & d** Magnified autofluorescence intensity and lifetime (LSC) corresponding to boxes in panel a. **e**, Mean (τ_m12_), short (τ_1_), and long (τ_2_) autofluorescence lifetimes within individual RPE cell (outlined by Voronoi polygons in panel d). **f** & **g**, Magnified autofluorescence intensity and lifetime (SSC) corresponding to boxes in panel b. **h**, Mean (τ_m12_), short (τ_1_), and long (τ_2_) lifetimes within individual RPE cells (Voronoi polygons in **g**). Corresponding multimodal retinal images are shown in **Extended Data Fig. 1**.

Inter-cellular variability, computed from per-cell median values within each region, were 6.66 ± 3.08 %, 13.09 ± 6.89 %, and 26.66 ± 13.27 % for τ_m12_, τ_1_, and τ_2_ in the LSC, respectively, and 9.24 ± 6.38 % and 49.55 ± 33.83 % for α_1_ and α_2_. In the SSC, rCV values were 11.00 ± 4.46 %, 22.48 ± 11.46 %, and 54.42 ± 28.35 % for τ_m12_, τ_1_, and τ_2_, and 18.94 ± 10.14 % and 97.37 ± 46.63 % for α_1_ and α_2_.

These results demonstrate pronounced intra-cellular and inter-cellular heterogeneity in fluorescence lifetime parameters at subcellular resolution, underscoring the capability of AOFLIO to resolve spatially structured lifetime variability within and between individual RPE cells in vivo.

Within 0°-10° eccentricity, the mean autofluorescence lifetime (τ_m12_) was 273.6 ± 53.2 ps in LSC and 211.3 ± 47.6 ps in SSC. For LSC, the short (τ_1_) and long (τ_2_) lifetime components were 188.3 ± 24.0 ps and 755.6 ± 220.7 ps, respectively, with corresponding relative amplitudes (α_1_ and α_2_, representing the wights of τ_1_ and τ_2_ in the total signal) of 82.7 ± 3.3 % and 15.2 ± 3.1%. SSC lifetimes were shorter, with τ_1_ = 154.5 ± 29.5 ps and τ_2_ = 591.1 ± 287.0 ps, corresponding α_1_ = 81.9 ± 3.7 % and α_2_ = 15.4 ± 6.1%.

### 2.3. AOFLIO Revealed the Effects of Age, Retinal Location, Gender, and Race/Ethnicity on RPE Autofluorescence Lifetime and Cellular Structure

Across fluorescence lifetime outcomes in both spectral channels, linear mixed-effects (LME) models analysis indicated that age and retinal eccentricity were the dominant sources of systematic variation. Mean lifetime (τ_m12_), short lifetime (τ_1_), and long lifetime (τ_2_) in both the LSC and SSC showed significant positive associations with age and retinal eccentricity (all p < 0.01; **Figs. 4a & 4b**; **Extended Data Table 1**). In contrast, amplitude parameters (α_1_ and α_2_) exhibited weaker and outcome-dependent associations, with significant age- or location-related effects observed for selected metrics only.

**Fig. 4.**
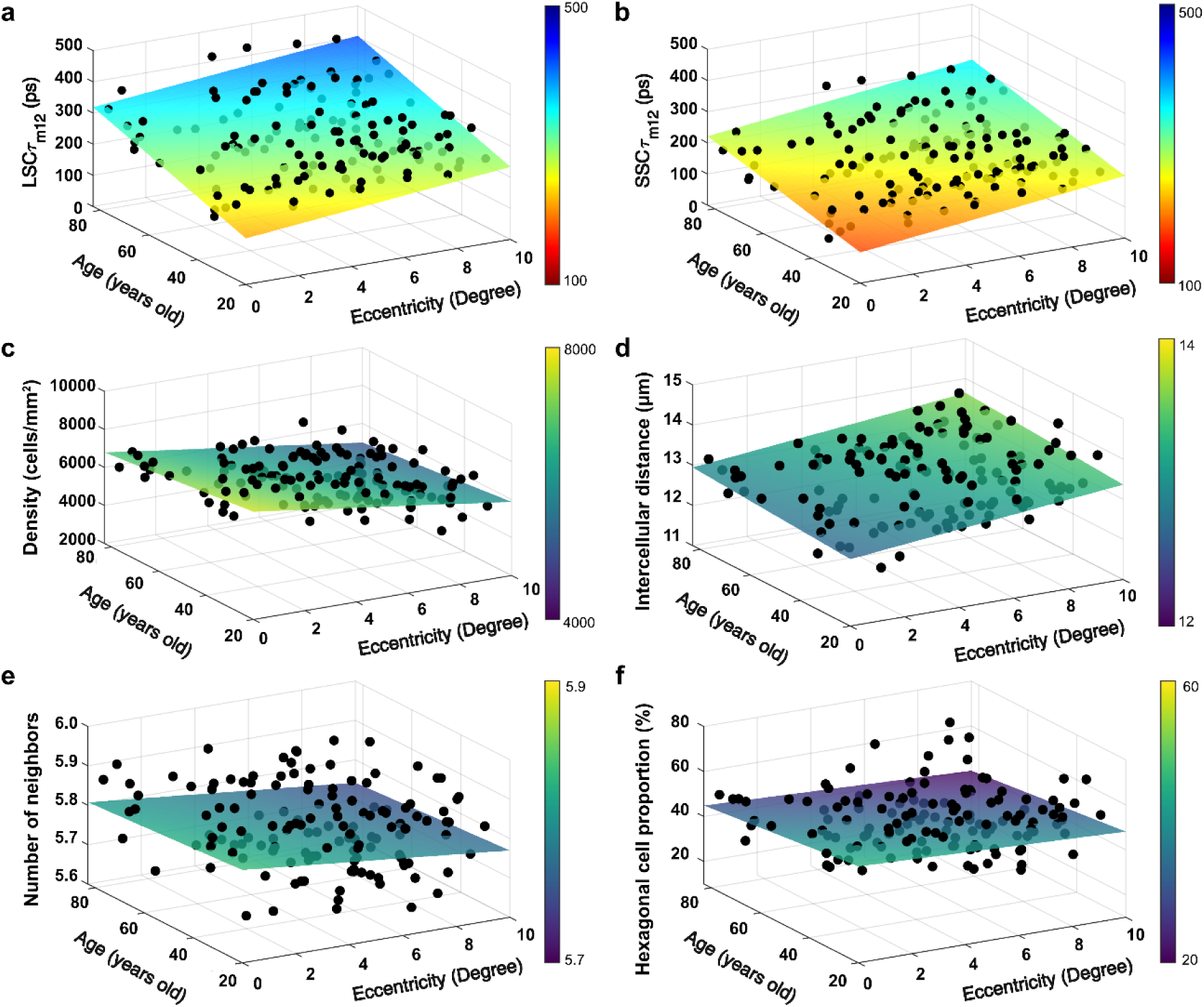
Normative distribution of retinal pigement epithelium (RPE) autofluorescence lifetime and cellular structural metrics as a function of age and retinal location. The corresponding model-fitted mean and 95% prediction confidence intervals that define the expected range of variability are shown in **Extended Data Fig. 3. a**, Mean RPE autoffluorescence lifetime in the long spectral channel. **b,** Mean RPE autoffluorescence lifetime in the short spectral channel. **c**, RPE cell density. **d**, Intercellular distance. **e**, Number of nearest neighboring cells. **f,** Proportion of hexagonal RPE cells.

For cellular structural measures, spatial variation was dominated by retinal eccentricity. RPE cell density decreased significantly with increasing eccentricity, whereas intercellular distance and the number of nearest neighboring cells increased with eccentricity (**Figs. 4c - 4e**; **Extended Data Table 1**). The proportion of hexagonal cells exhibited weaker associations with both age and eccentricity and did not show a consistent monotonic trend across the sampled retinal locations (**Fig. 4f**).

Gender and race/ethnicity contributed minimally to model fit for most lifetime and structural outcomes **(Extended Data Table 1)**. Likelihood ratio tests in full models showed that removal of either term did not significantly degrade model fit after false discovery rate correction (p > 0.05, **Supplementary Table 7 – 9**).

Based on systematic ‘drop-one’ model comparisons, a reduced core mixed-effects model including age and retinal eccentricity was identified as a parsimonious representation of normative variation across outcomes (**Extended Data Table 2**). This core model was therefore adopted to define a low-dimensional normative distribution of RPE autofluorescence lifetime and cellular structural metrics as a function of age and retinal location. This model incorporated both between-subject variability and within-subject residual variance, thereby defining normative cellular-level variability and providing a quantitative reference for assessing deviations from the healthy distribution in disease conditions (**Fig. 4**; **Extended Data Fig. 3**).

### 2.4 AOFLIO Revealed Cellular-Scale RPE Metabolic and Morphometric Alterations in AMD

AMD is a leading cause of central vision loss in the developed countries and worldwide. It is a multifactorial disease involving the photoreceptors, RPE, Bruch’s membrane (BrM), and choroidal vasculature.^56^ Although the events that give rise to retina-RPE-choroid pathology in AMD remain incompletely understood, a major early hallmark is the accumulation of extracellular pathological material either in the sub-RPE space between the RPE and the inner surface of Bruch’s membrane (BrM), forming drusen,^57,58^ or in the subretinal space between the RPE and photoreceptors, forming subretinal drusenoid deposits (SDDs).^59–61^ These lesions separate photoreceptors from their blood supply and promote RPE and outer retinal atrophy and neovascularization, causing significant visual impairment.^56^ Thus, RPE is a critical AMD participant, victim, and a reporter of clinically invisible events in the BrM and a target for therapy.^56,62^ Understanding the functional and structural changes in the RPE during aging and AMD progression can help identify early biomarkers and therapeutic targets.

High-resolution AOFLIO demonstrated lesion-type–dependent deviations in RPE metabolic function and structure in regions containing AMD lesions (**Fig. 5**; **Supplementary Figs. 3–4**). Compared with healthy controls, RPE autofluorescence lifetimes in drusen and SDD regions exhibited increased τ_m12_ along with coordinated changes in lifetime components and amplitudes across both channels (**Figs. 5c, 5d, 5i, 5j**). Spatially resolved phasor maps delineated distinct functional boundaries aligned with lesion morphology (**Figs. 5e, 5f, 5k, 5l**) and revealed high resolution functional gradients in advanced pathology, such as collapsing drusen and geographic atrophy borders (**Fig. 6; Supplementary Figs. 5–6**). These features were not apparent in intensity images or in clinical FLIO.

**Fig. 5.**
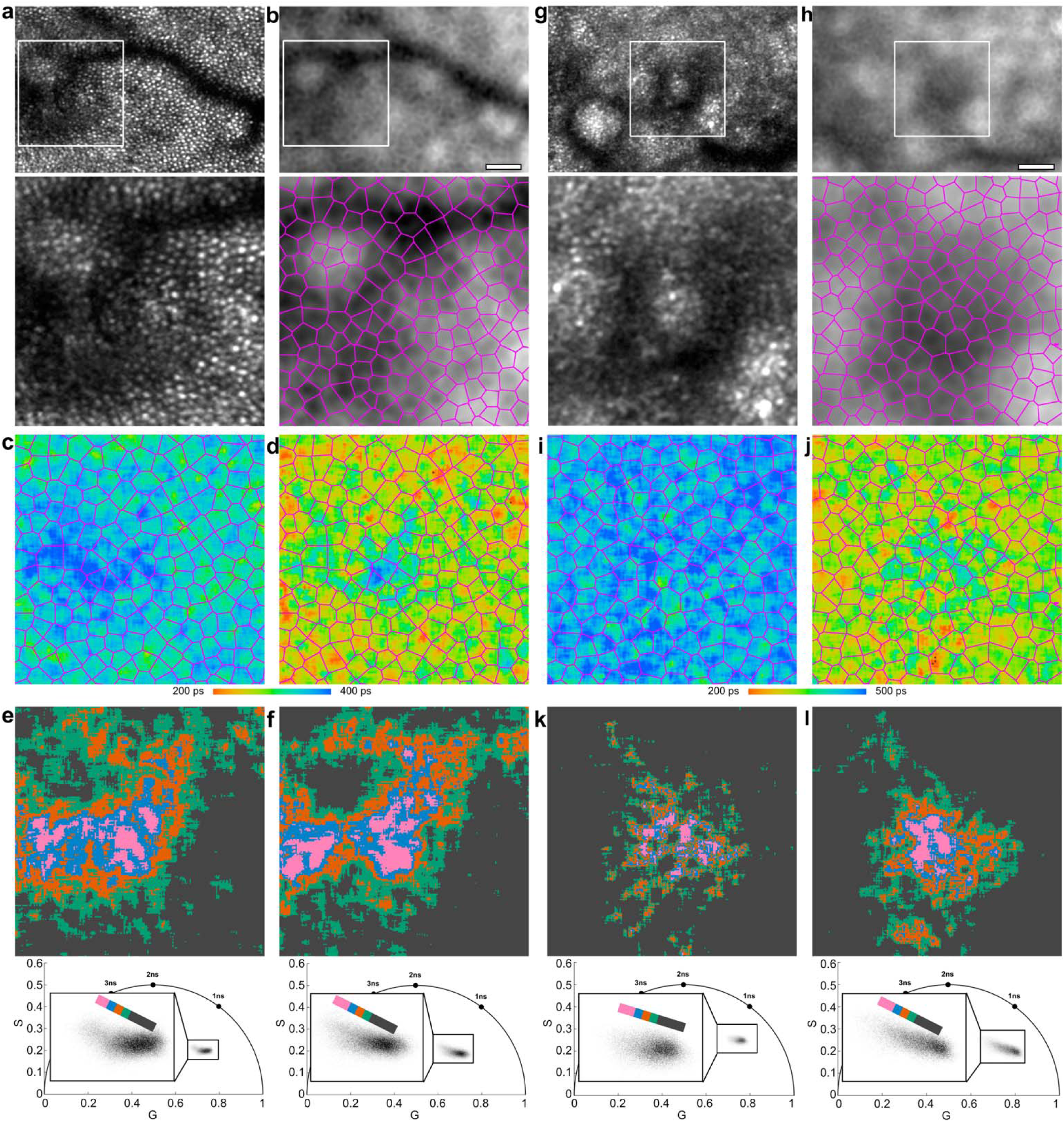
Adaptive optics fluorescence lifetime imaging ophthalmoscopy (AOFLIO) resolves cellular-scale functional alterations of the retinal pigment epithelium (RPE) in eyes with age-related macular degeneration (AMD). **a–f**, RPE autofluorescence in a region containing a druse. **a**, AOSLO reflectance image (λ = 790 nm) showing photoreceptors overlying a large druse. Scale bar, 50 µm. **b**, Autofluorescence photon intensity image with RPE cells delineated using Voronoi tessellation. **c & d,** RPE autofluorescence lifetime maps acquired in the long spectral channel (LSC) and short spectral channel (SSC), respectively. **e,f**, Autofluorescence lifetime images are color-coded by phasor analysis results in the LSC and SSC, respectively, delineating lifetime differences in RPE overlying the druse with cellular resolution. **g–l**, RPE autofluorescence in a region containing a subretinal drusenoid deposit (SDD), shown using the same panel order and representations as in a–f. Corresponding clinical multimodal images characterizing these lesions are shown in **Supplementary** Figs. 3–4.

**Fig. 6.**
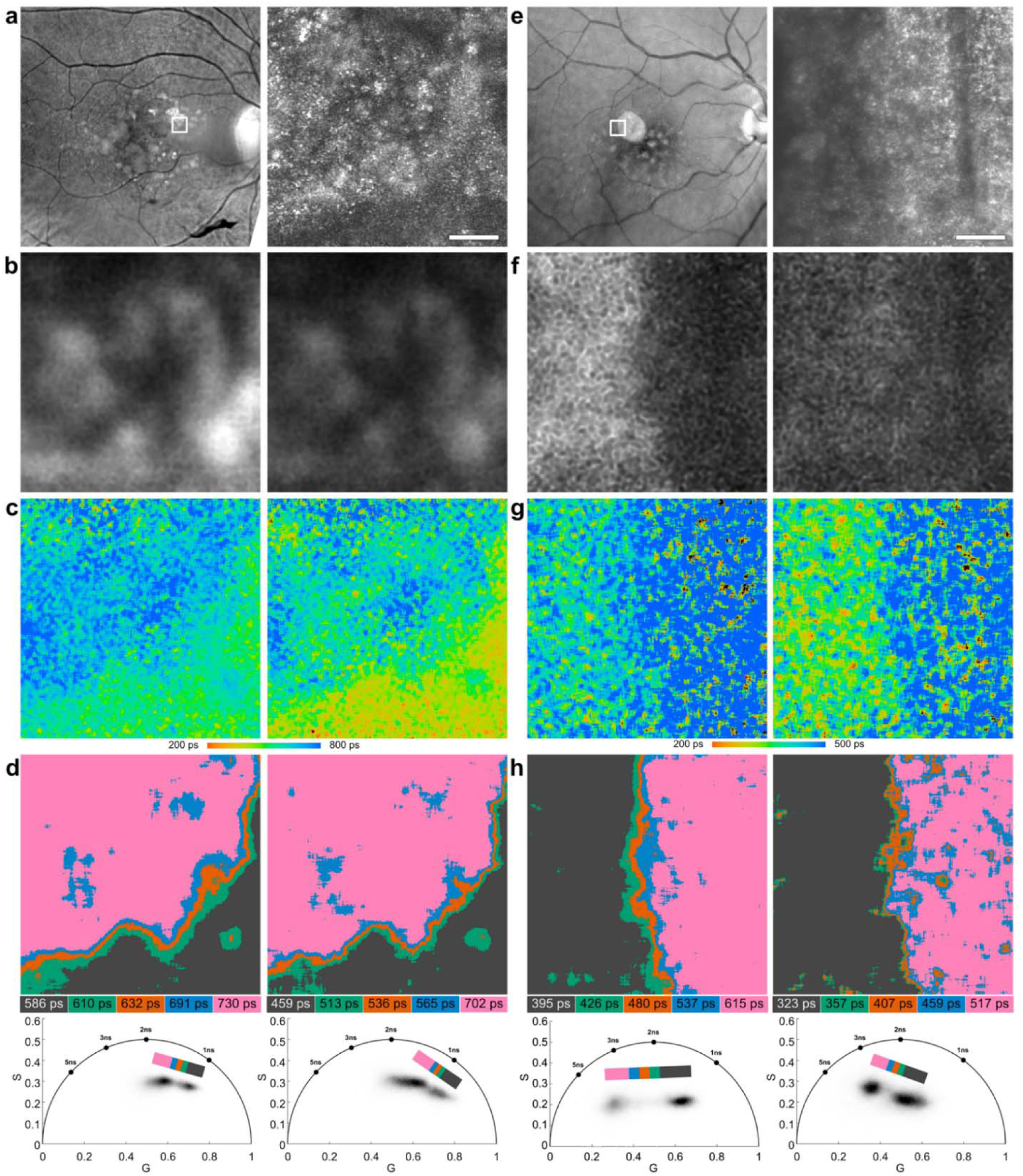
Adaptive optics fluorescence lifetime imaging ophthalmoscopy (AOFLIO) delineates functional boundaries and transition zones in advanced age-related macular degeneration (AMD) lesions at cellular-scale resolution. Panels **a–d** show an example of a collapsing druse. **a**, En face near-infrared reflectance image showing a collapsing druse (white box) and the corresponding high-resolution AOSLO reflectance image (λ = 790 nm). Scale bar, 100 µm for AOSLO and AOFLIO images in panels a–d. **b**, Autofluorescence photon intensity images acquired in the long (left) and short (right) spectral channels. **c**, Corresponding autofluorescence lifetime maps. **d**, spatially resolved phasor analysis reveals heterogeneous lifetime signatures and highlights functional boundaries and transitions at the lesion edge. Panels **e–h** present a case of geographic atrophy (GA), shown using the same panel order and representations as in panels a–d. Phasor-based visualization provides a fine delineation of the functional boundaries in regions adjacent to the atrophic area. Corresponding clinical multimodal imaging characterizing these lesions is shown in **Supplementary** Figs. 5–6.

Group-based mixed-effects modeling across AMD lesion types, including drusen,^63^ SDD,^64^ basal laminar deposit (BLamD),^65^ acquired vitelliform lesions (AVL),^66^ and geographic atrophy (GA),^67^ revealed significant RPE metabolic and structural differences between eyes with AMD and healthy controls, including in clinically unremarkable retinal regions. After accounting for retinal eccentricity and within-subject correlation, RPE cells in clinically unremarkable regions of AMD eyes exhibited significantly prolonged mean autofluorescence lifetimes (τ_m12_) compared to age-matched controls in both spectral channels (LSC, β = 72.96, p = 0.0009; SSC, β = 39.45, p = 0.0308; **Extended Data Tables 3**), accompanied by systematic shifts in lifetime components and amplitude fractions, indicating a global RPE metabolic alteration in AMD extending beyond regions with clinically apparent lesions.

Compared to unremarkable regions, RPE autofluorescence lifetimes in drusen and SDD areas showed further significant increases in SSC τ_m12_ (drusen: β = 19.01, p = 0.0271; SDD: β = 23.35, p = 0.0212), whereas LSC τ_m12_ differences were smaller and not consistently significant (drusen: β = 14.42, p = 0.0730; SDD: β = 8.85, p = 0.3462; **Extended Data Table 4**), suggesting lesion-specific metabolic perturbations superimposed on a broader AMD background.

Structural metrics also differed between AMD and healthy eyes. In unremarkable regions, RPE cell density was lower (β = −604.73, p = 0.0033) and intercellular distance was higher compared with healthy controls (β = 1.48, p < 0.0001), while hexagonality did not differ significantly (**Extended Data Table 3**). Within AMD eyes, comparisons with unremarkable regions showed significant differences in RPE cell density, intercellular distance, and nearest-neighbor counts for SDD regions, and in number of neighbors for drusen regions (**Extended Data Table 4**).

## 3. Discussion

In this work, we developed AOFLIO, enabling single-cell–resolved in vivo metabolic and structural imaging of the human RPE by overcoming key technical barriers to cellular-level fluorescence lifetime imaging in the living eye. A central advance is the direct reconstruction of RPE cellular structure from the same autofluorescence photons captured for lifetime measurements, providing intrinsic structure–function co-registration at the single-cell level. AOFLIO achieves robust cellular resolution in both long and short spectral channels, including the photon-limited short spectral channel sensitive to FAD-related metabolic activity.

A critical challenge for cellular-level fluorescence lifetime imaging in the living human retina has been the lack of a practical and accurate mechanism to register sparse autofluorescence photons with sufficient spatial precision. This limitation has impeded direct resolution of individual RPE cell structure from the photons captured for lifetime measurement. We addressed this challenge through a highly accurate timing architecture that enforces deterministic phase synchronization between pilot AOSLO reflectance imaging and TCSPC acquisition of RPE autofluorescence photons. As a result, each detected photon is assigned a precise temporal timestamp corresponding to a definitive spatial coordinate of its origin.

In a multi-wavelength system, chromatic aberration introduces additional challenges.^68–71^ In multi-wavelength imaging systems, chromatic aberration introduces additional challenges. In addition to optical mitigation of both lateral and longitudinal chromatic aberrations in the living human eye, we further reduce residual lateral chromatic offsets among spectral channels through exact spectra-specific, independently calibrated confocal detection. This strategy enables accurate motion correction across spectral channels and intrinsic high-precision registration between excitation reflectance and autofluorescence imaging. High-frequency TCSPC further enables subpixel subdivision of the pilot AOSLO reflectance image pixels, allowing autofluorescence photons to be localized beyond the native AOSLO sampling grid. Non-interpolative linearization of sinusoidal distortion in both AOSLO and AOFLIO images minimizes temporal ambiguity in photon assignment that would otherwise arise from interpolative redistribution of photon events, which assigns the same micro-time to multiple spatial locations and introduces uncertainty in autofluorescence decay estimation.^72^

Collectively, these advances distinguish the present work from prior AO-assisted fluorescence lifetime imaging approaches^38–41,72^ and enable accurate assignment and registration of autofluorescence photons across time, frames, and spectral channels. This capability yields reliable per-cell fluorescence decay estimates and makes single-cell–resolved AOFLIO feasible in vivo.

Notably, this performance arises not from incremental improvements in optical resolution or photon throughput, but from a fundamentally new photon-level temporal–spatial encoding and reconstruction strategy.

Single-cell-resolved AOFLIO enabled quantification of intra- and inter-cellular autofluorescence lifetime variability and the establishment of normative single cell autofluorescence lifetime benchmarks, revealing systematic increases with age and retinal eccentricity. Morphometric analysis confirmed eccentricity-dependent gradients in cell density, spacing, and hexagonality, consistent with histologic measurements^14,18,73^ and prior AO structural imaging studies.^9,74–83^ Importantly, In healthy eyes, per-cell AOFLIO lifetime analysis demonstrated age- and eccentricity-dependent increases in the mean lifetime τ_m12_ and its components (τ_1_, τ_2_) in both spectral channels. Because these trends were derived from subcellular autofluorescence decay rather than spatially averaged regions, they provide quantitative estimates of the RPE functional state that reflect cell-intrinsic fluorophore composition and microenvironmental influences not accessible through structural imaging alone.

In the LSC, age-related increases in the slow-component fraction (α2) and prolongation of τ1 and τ2 are consistent with progressive accumulation of long-lived bisretinoid lipofuscin and melanolipofuscin within individual RPE cells and known eccentricity-dependent granule loading (**Extended Data Table 1**). These relationships, previously inferred largely from histology and areal measurements,^4,49,84^ are here demonstrated at the single-cell level. Although α1 and α2 remained relatively stable across eccentricity, the concomitant lengthening of τ1 and τ2 indicates position-dependent intracellular modulation of autofluorescence lifetime determinants, including viscosity, refractive index, redox/oxygen quenching, and non-radiative decay pathways.^22,85,86^. In the SSC, cell-resolved increases in τ_m12_, τ_1_, and τ_2_ are consistent with a composite signal dominated by oxidized flavins whose lifetimes are sensitive to binding state and microenvironment, superimposed on the very short-lived macular pigment contribution in the central macula.^10,87^ The absence of race- or sex-related effects suggests these cell-level age- and eccentricity-dependent signatures suggests that these signatures reflect generalizable RPE physiology, thereby establishing a normative cellular baseline for detecting metabolic alterations in disease.

AOFLIO was further applied to studying AMD, demonstrating its applicability under pathological conditions. Marked RPE autofluorescence lifetime alterations were detected in regions with drusen, SDD, BlamD, AVL, and GA, as well as in clinically unremarkable areas (**Figs. 5 & 6, Extended Data Fig. 4**), indicating the presence of subclinical dysfunction. These functional abnormalities were accompanied by structural remodeling of the RPE mosaic, including reduced cell density, increased spacing, and altered hexagonality, which closely paralleled these functional changes. Lesion-specific fluorescence lifetime signatures delineated distinct pathological states, highlighting the sensitivity of AOFLIO to disease-relevant metabolic heterogeneity that is inaccessible to conventional imaging.

Although these observations were obtained from a relatively small number of subjects, the consistency of lesion-specific functional alterations, together with their associated structural changes, provide plausible clinical hypotheses and mechanistic frameworks motivating future investigation larger, well-powered cohorts.

This study has important limitations. AOFLIO was implemented with a relatively large confocal pinhole (150 µm, corresponding to 5.0 Airy disk diameters (ADD) in the LSC and 6.3 ADD in the SSC) to satisfy photon collection under light-safety constraints, reducing axial sectioning (∼70 µm). The current photon budget required 9 × 9-pixel binning (∼6.0 µm) for reliable lifetime estimation, limiting the full exploitation of AO spatial resolution, and the acquisition time (∼100 s) remains longer than ideal for routine clinical application. Future technical advances, including advanced phasor analysis^88^ and machine-learning–based lifetime estimation under low-photon conditions,^89,90^ are expected to reduce pinhole size, minimize binning requirements, shorten acquisition time, and improve clinical feasibility.

Beyond imaging RPE metabolism, with further improvements in confocal performance and axial selectivity particularly in the 500–530 nm emission band under 473 nm excitation where flavin-based fluorescence is prominent,^91^ AOFLIO may enable in vivo metabolic imaging of inner retinal neurons. Such capabilities may provide a window into neurodegenerative conditions in which early metabolic dysfunction is implicated, such as glaucoma, mitochondrial optic neuropathies, and neurodegenerative disorders with retinal involvement, including Alzheimer’s and Parkinson’s disease.^92^

In conclusion, by integrating adaptive optics-enabled cellular resolution with fluorescence lifetime imaging, a nascent modality offering molecular contrast, AOFLIO overcomes the limited spatial resolution of conventional FLIO and the predominantly structural focus of existing adaptive optics retinal imaging. This integration enables high-resolution functional “optical biopsy” at the cellular and subcellular levels, where dysfunction often precedes structural degeneration, and provides objective metabolic biomarkers for assessing disease risk, progression, and potentially therapeutic response in retinal and neurodegenerative conditions. These advances establish AOFLIO as a robust and transformative platform for cell-resolved functional and structural imaging, expanding the ophthalmic imaging landscape to a previously inaccessible biological scale in vivo.

## 4 Methods

### 4.1 Compliance, Human subjects, and Standard Clinical Characterization

This study adhered to the tenets of the Declaration of Helsinki, complied with the Health Insurance Portability and Accountability Act of 1996, and was approved by the Institutional Review Boards at the University of California, Los Angeles. Written informed consent was obtained from all participants prior to imaging. Human subjects, including normal controls and patients with AMD, were recruited from the clinical research registry of the Doheny Eye Center, University of California - Los Angeles. All participants underwent comprehensive ophthalmic examination supplemented by multimodal clinical imaging to characterize retinal health and pathology. Detailed inclusion and exclusion criteria, subject demographics, and clinical imaging protocols are provided in **Supplementary Note 1**.

### 4.2 AOFLIO Imaging

Eligible subjects were invited to participate in AOFLIO imaging. Before imaging, the subject’s pupil was pharmacologically dilated using 1.0% tropicamide and 2.5% phenylephrine hydrochloride. Head position was aligned and stabilized with a head mount and chin rest. Fixation was guided by a flashing bright cross displayed on a screen. During acquisition, the fixation target was repositioned to control the subject’s viewing angle. At each retinal location, 1,500 frames were acquired over 100 seconds. Videos were recorded at 5–6 locations along the primary nasal retinal meridian extending to 10° eccentricity. A complete AOFLIO imaging session lasted 20-30 minutes, including rest periods. Prior to data acquisition, the gains of the AOSLO and AOFLIO photodetectors were adjusted to achieve appropriate image brightness and contrast based on real-time histograms of retinal videos and were held constant throughout the imaging session.

### 4.3 Retinal Autofluorescence Photon Temporal-Spatial Reassignment and Registration

#### 4.3.1 Pixel consolidation based non-interpolation desinusoidal correction

Sinusoidal scan distortion from the resonant mirror in AOFLIO was corrected using a non-interpolative desinusoidal remapping approach (**Extended Data Fig. 1**). Unlike conventional image linearization, which interpolates pixel intensities and split individual photons.^72^ AOFLIO pixels consist of discrete photon events carrying micro-time stamps for fluorescence lifetime estimation and therefore cannot be fractionally redistributed. Photon events were reassigned as indivisible units to corrected spatial origins.

Scan velocity was calibrated for each session using a custom grid imaged at the retinal plane across all detection channels (AOSLO reflectance at 473 nm and 790 nm; AOFLIO LSC and SSC). For each scan line, pixel spatial sizes were estimated from grid spacing, and the pixel at peak scan velocity was used as a reference. Adjacent pixels acquired at lower velocities were consolidated into a single corrected pixel such that their combined physical size matched the reference, using a sliding consolidation rule. This generated a spatiotemporal lookup table converting sinusoidal temporal sampling into uniform spatial sampling without interpolation.

Because the AOFLIO pixel clock operated at four times the AOSLO rate, photons were localized with one-quarter–pixel precision (≈0.17 µm). Channel-specific calibration minimized residual chromatic misregistration, and forward and backward scans were merged to increase photon counts while preserving lifetime information.

#### 4.3.2 Two-stage substrip-based high-precision photon registration

To construct autofluorescence decay curves at each AOFLIO pixel, individual photons must be accurately registered to their spatial origins. Because autofluorescence photons are sparse, direct cross-correlation–based registration is not feasible. Instead, retinal motion was estimated from simultaneously acquired, high-contrast AOSLO photoreceptor images and used to guide photon registration. A two-stage substrip-based registration strategy was implemented to minimize motion-induced and chromatic misregistration.

In the first stage, simultaneously acquired AOSLO reflectance images (790 nm and 473 nm) and AOFLIO photon intensity images (LSC and SSC) were segmented into temporal groups defined by microsaccade boundaries, as determined from the 790 nm AOSLO images. Each group spanned approximately 30–100 frames. Within each group, eye motion was estimated using substrip-based registration of the 790 nm AOSLO images. The resulting motion vectors were applied to the corresponding 473 nm AOSLO images and AOFLIO photon data to correct intra-group motion,, yielding a registered 473 nm AOSLO image for each group.

In the second stage, the motion-corrected 473 nm AOSLO images from all groups were registered across the full acquisition period (∼100 s) to estimate inter-group motion, which was subsequently applied to the AOFLIO photon data. Following registration, photons were assigned to AOFLIO pixels and sorted into per-pixel arrival-time histograms, enabling fluorescence decay reconstruction and lifetime fitting.

### 4.4 Fluorescence Lifetime Analysis

#### 4.4.1 Model based autofluorescence lifetime calculation

Fluorescence lifetime analysis was performed using commercial software (SPCImage, Becker & Hickl GmbH, Germany). The fluorescence decay was modeled using a tri-exponential function that accounted for the early arrival of human lens autofluorescence,^93^

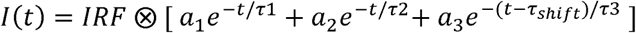

where I (t) denotes the fluorescence intensity and IRF is the instrument response function, *a_1_*, *a_2_*, and *a_3_* are the weight of each component to the total fluorescence and determined by the concentration of a fluorophore.<u>94</u> r_shift_ is a time shift accounting for the early arrival of lens autofluorescence. The mean RPE autofluorescence lifetime was calculated by, 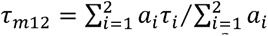. The weight of lifetime component (*i*) in the total fluorescence was calculated by 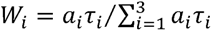.

To ensure accurate fitting, spatial binning of 9 × 9 pixels was applied, with a minimum threshold of 1000 photons per decay curve. The *IRF* was modeled using the manufacturer-recommended parametric function *t/t_w_*·exp(*–t/t_w_*), where t denotes the photon arrival time relative to the excitation pulse and t_w_ is a width parameter describing the temporal dispersion of the detection system. The *IRF* position was automatically aligned to the experimental decay by SPCImage. Fluorescence decay curves were then convolved with this IRF and fitted with the tri-exponential model using nonlinear least-squares minimization.

#### 4.4.2 Phasor analysis

Fluorescence lifetime was additionally analyzed using a phasor-based approach, which provides a fit-free, frequency-domain representation of fluorescence decays. For each pixel, the fluorescence decay I(t) was transformed into phasor coordinates (G, S) at the laser repetition frequency:

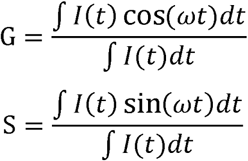

where ω = 2πf and f is the laser repetition rate. Measured phasors were corrected for the IRF using a parametric IRF model consistent with the model-based lifetime analysis. Phasor distributions were visualized as density histograms and color-coded scatter plots, and phasor-based classifications were mapped back to image space to generate spatially resolved lifetime maps.

### 4.5 Inter-channel Correlation of RPE Cell Structure

Pixel-wise Pearson correlation coefficients (r) between SSC and LSC intensity images were computed to assess spatial concordance of autofluorescence signals across AOFLIO channels. Intensity images were generated by summing registered autofluorescence photons, with no additional smoothing, thresholding, or outlier rejection; all valid pixels were included.

For each retinal location, Pearson’s coefficient was calculated across all valid pixels, yielding a single correlation value per location. This analysis was performed at 152 retinal locations from 34 healthy eyes. Correlation coefficients are reported as mean ± SD with corresponding 95% confidence intervals. Statistical significance relative to zero was assessed using a two-sided one-sample t-test.

### 4.6 RPE Cellular Morphology Analysis

AOFLIO intensity images were median filtered to reduce photon-count noise. RPE cell centers were manually identified as dark nuclear profiles using ImageJ (v1.53t, National Institutes of Health, USA). These coordinates were used to generate Voronoi tessellations, delineating individual RPE cell boundaries. Cellular structure was analyzed using a custom MATLAB script.^95^ Metrics included RPE cell density, nearest neighbor spacing, intercellular distance, neighbor count distribution, and the proportion of hexagonal cell. All metrics were computed consistently across healthy controls and AMD eyes and used as inputs for subsequent spatial and group-based analyses.

### 4.7 Analysis of Intra- and Inter-cellular Variability of RPE Autofluorescence Lifetime

To quantify cellular-scale variability in RPE autofluorescence lifetime, individual RPE cells were segmented using Voronoi tessellations derived from AOFLIO intensity images, and these boundaries were applied to the corresponding lifetime maps. Lifetime values were extracted on a pixel-wise basis within each cell.

Because within-cell lifetime distributions were often non-Gaussian, variability was quantified using a robust coefficient of variation (rCV), defined as the interquartile range (IQR) divided by the median.

Intra-cellular variability was calculated as the rCV of pixel-wise lifetimes within each cell, and the distribution of these values was summarized across cells. Inter-cellular variability was assessed by computing the median lifetime for each cell and then calculating the rCV across these cellular medians, providing a measure of variability among cells.

### 4.8 Modeling RPE Autofluorescence Lifetime and Cellular Structure in Normal Healthy Eyes

Associations between RPE autofluorescence lifetime parameters and cellular structural metrics with age, retinal eccentricity, sex, and race were assessed using linear mixed-effects models with a subject-specific random intercept to account for repeated retinal measurements. Age and eccentricity were treated as continuous variables and standardized prior to analysis. Models were fit using maximum likelihood. Fixed effects were evaluated using drop-one likelihood ratio tests, with model comparison guided by likelihood ratio tests and information criteria (AIC and BIC) and false discovery rate control across outcomes. A reduced core model including age and eccentricity was used to define a normative reference distribution for healthy participants (**Fig. 4; Extended Data Fig. 3**). Models were implemented in MATLAB (Statistics and Machine Learning Toolbox) using standard mixed-effects modeling functions.

### 4.9 Group-based mixed-effects analysis of cellular-scale AOFLIO outcomes in AMD

AMD lesion types and unremarkable retinal regions were defined using standardized multimodal imaging criteria (**Supplementary Note 1**) and localized in AOFLIO images by co-registering clinical OCT with AOSLO reflectance images (**Figs. 5 and 6, Supplementary Note 5**). RPE cellular segmentation and morphometric analysis were performed as described in **Section 4.6**. Spatial heterogeneity of autofluorescence lifetime was additionally visualized using phasor analysis to delineate functional boundaries without assuming a decay model.

Group-level differences in RPE autofluorescence lifetime parameters (τm12, τ1, τ2, α1, α2 in both channels) and cellular structural metrics were analyzed using linear mixed-effects models to account for repeated measurements within individuals. Group membership (healthy controls, AMD unremarkable regions, and lesion-specific regions) was included as a categorical fixed effect, and retinal eccentricity was included as a standardized continuous covariate. All models included a subject-specific random intercept. Healthy controls (≥60 years) were age-matched to the AMD cohort.

Overall group effects were assessed using Type III tests. When significant, post hoc contrasts compared unremarkable regions versus controls, lesion regions versus controls, and lesion regions versus unremarkable regions, distinguishing global AMD-associated changes from lesion-specific effects. Models were fit using restricted maximum likelihood, and effect estimates are reported with confidence intervals and p-values.

## Extended Data

**Extended Data Fig. 1.**
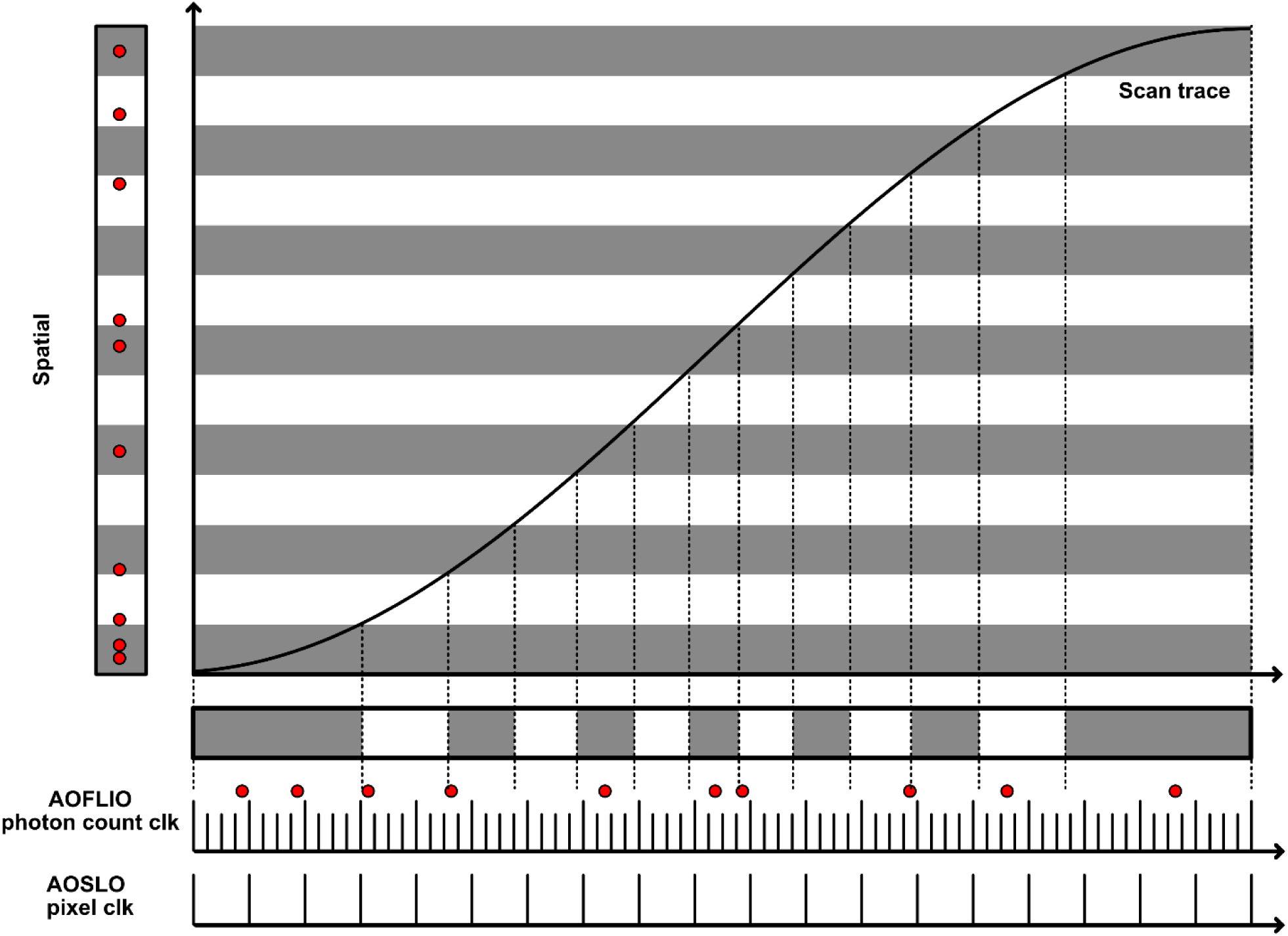
High-precision temporal–spatial mapping for photon reassignment in AOFLIO. Autofluorescence photons are acquired along a sinusoidally distorted scan trajectory due to resonant horizontal scanning. To correct this nonlinearity, a custom fluorescent calibration grid was imaged to measure the scanning velocity profile and establish a deterministic temporal–spatial mapping. Each imaging channel was calibrated independently to minimize residual ocular chromatic aberration. Using this mapping, individual photon events were treated as indivisible units and reassigned to their corrected spatial coordinates via non-interpolative pixel consolidation rather than intensity interpolation. Because the AOFLIO pixel clock operates at four times the AOSLO rate, each AOSLO reflectance pixel was virtually subdivided into four AOFLIO subpixels, enabling photon localization with one-quarter–pixel spatial precision relative to AOSLO. This approach converts photons acquired at uniform temporal intervals into a spatially uniform representation while preserving photon statistics, fluorescence lifetime information, and effective spatial resolution.

**Extended Data Fig. 2.**
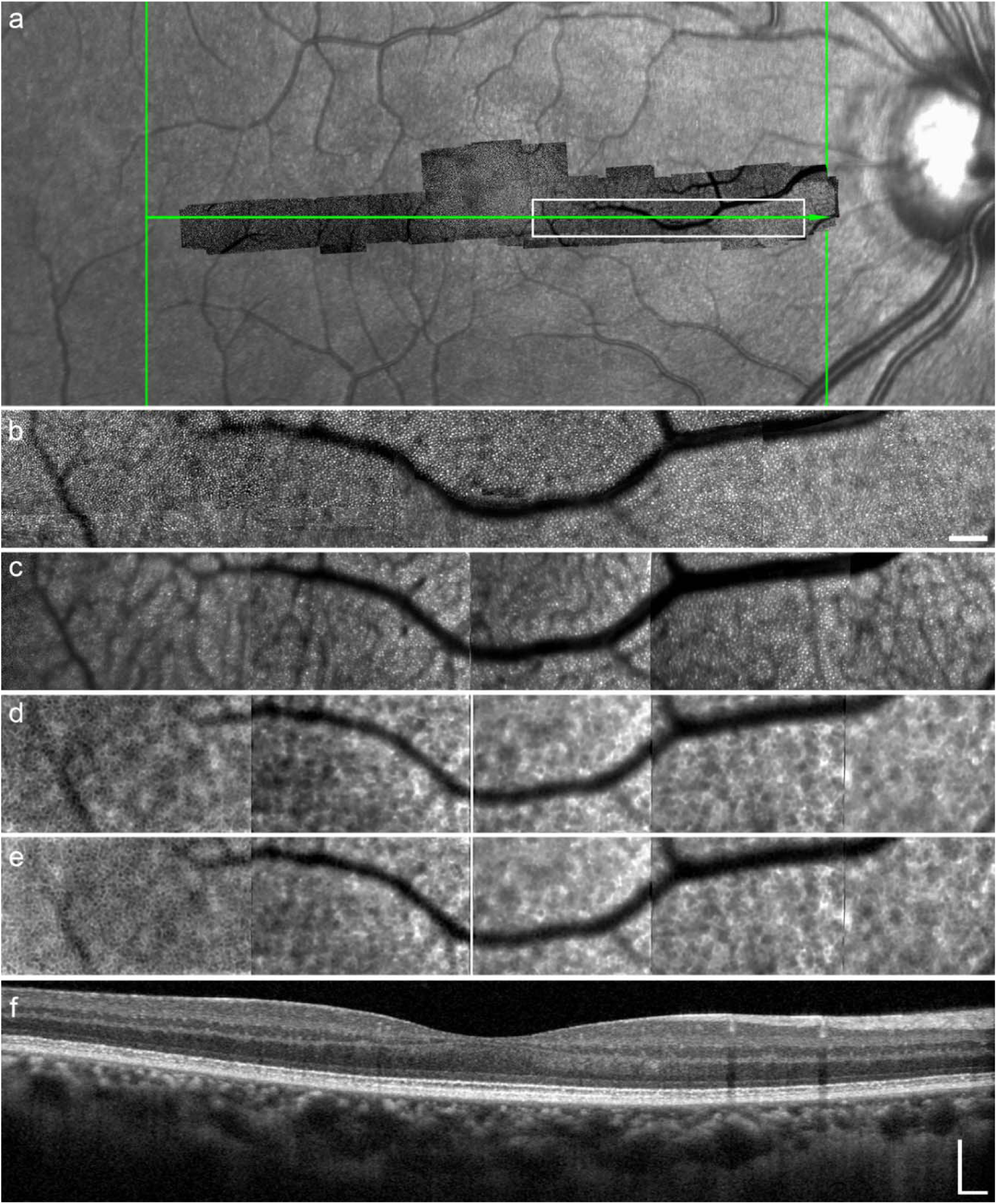
Multimodal retinal imaging corresponds to Fig. 3. a, Fundus photograph of the left eye with AOSLO imaging field overlaid. The white boxes indicate the regions shown in b–e, and the green line marks the location of the OCT B-scan shown in f. b, AOSLO reflectance image acquired at 790 nm. Scale bar, 100 µm. c, AOSLO reflectance image acquired at 473 nm. d, Autofluorescence (AF) intensity image in the long spectral channel (LSC, 560∼720 nm). e, AF intensity image in the short spectral channel (SSC, 497∼560 nm). f, OCT B-scan corresponding to the green line indicated in a. Scale bar, 200 µm.

**Extended Data Fig. 3.**
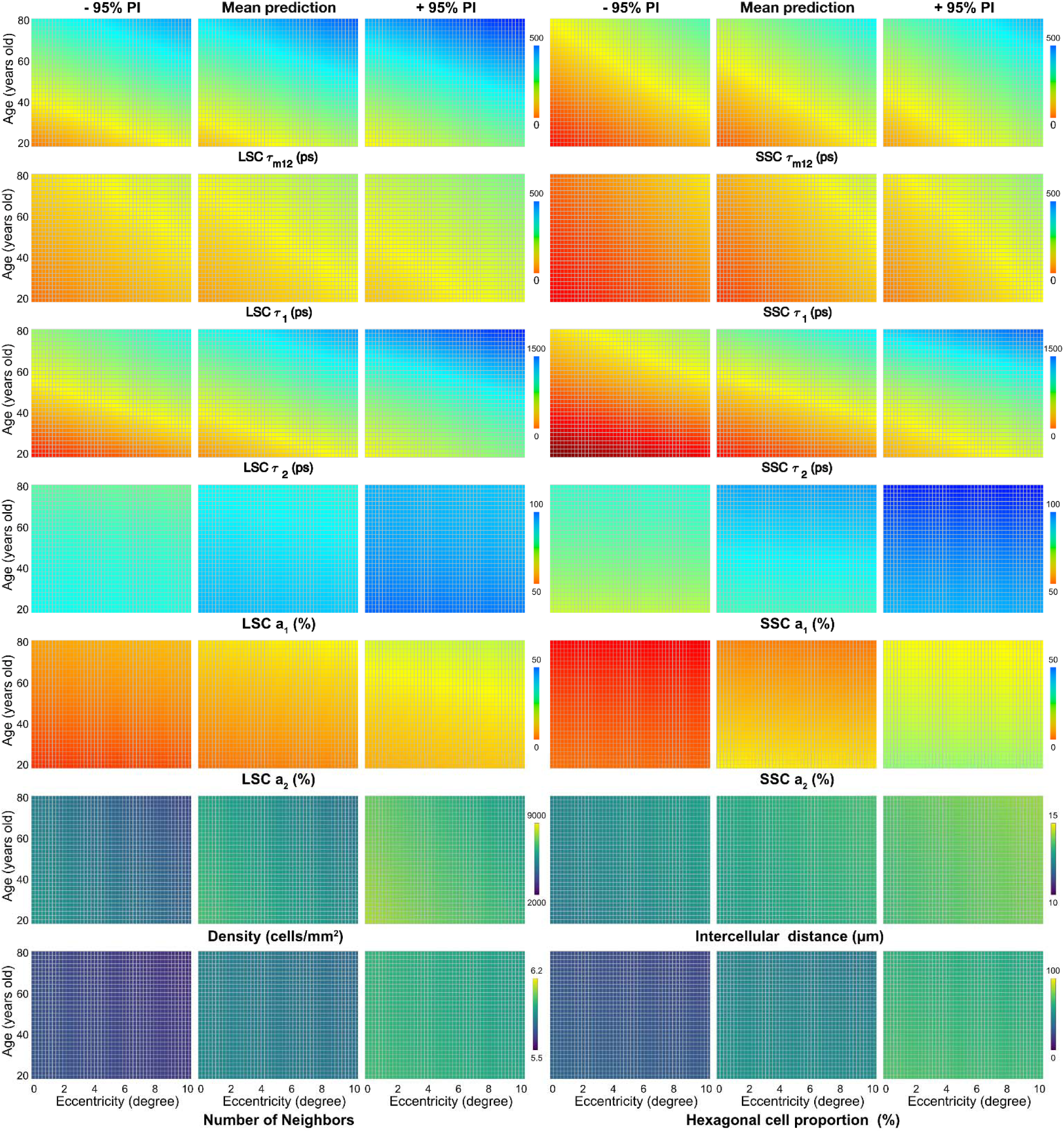
Normative distribution of retinal pigment epithelium (RPE) autofluorescence lifetime and cellular structural metrics as a function of age and retinal location. The mean and 95% confidence intervals were estimated using a linear mixed-effects model, defining a normative variability envelope that provides a quantitative reference for evaluating deviations from healthy cellular-level retinal structure and autofluorescence lifetime.

**Extended Data Fig. 4.**
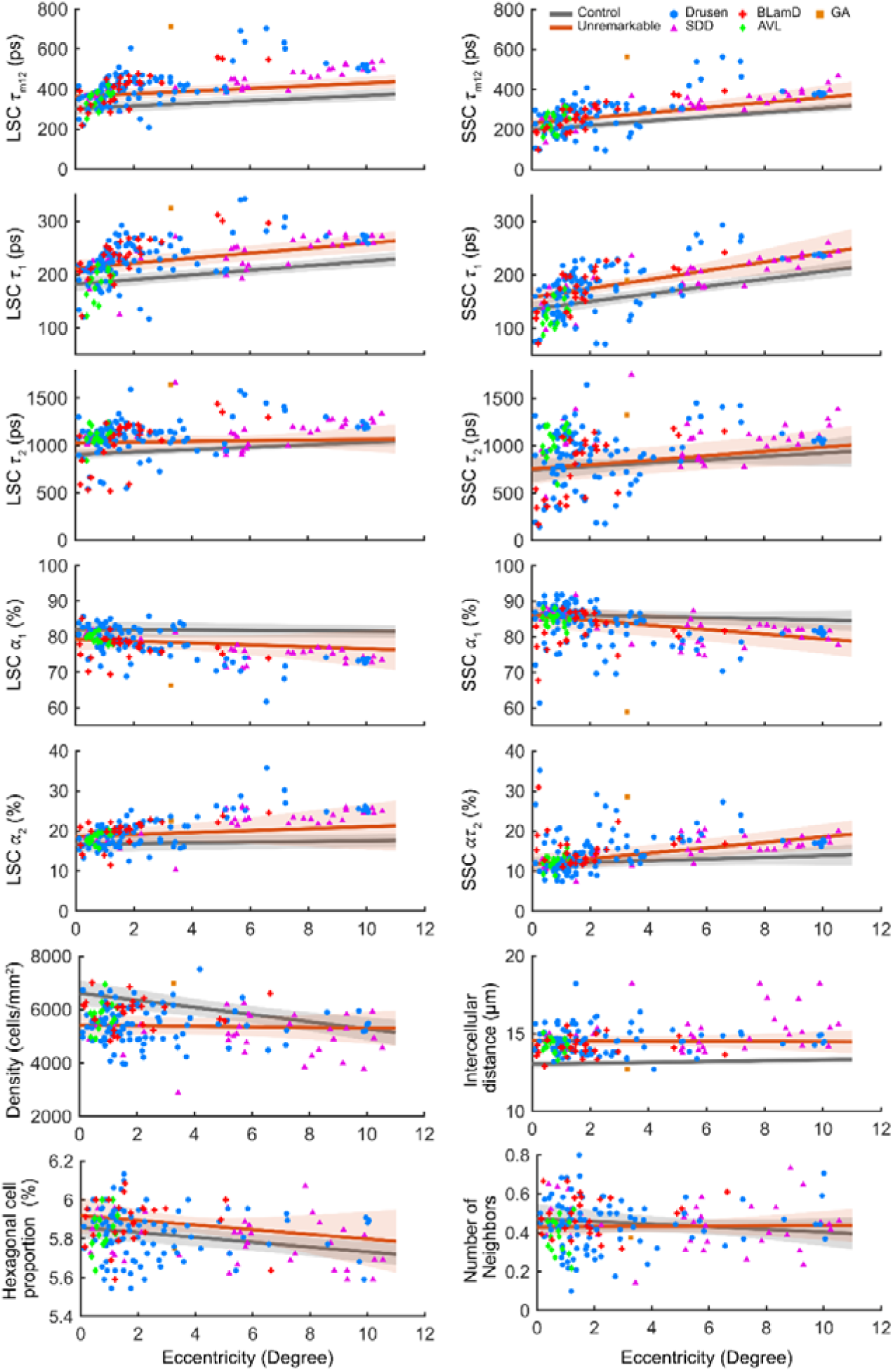
Location-dependent reference trends and lesion-associated measurements in AMD eyes. Location-dependent profiles of autofluorescence lifetime parameters (τ_m12_, τ_1_, τ_2_, α_1_, α_2_ in both channels) and cellular structural metrics (RPE cell density, intercellular distance, number of neighbors, and hexagonal cell proportion) are shown as functions of retinal eccentricity. For each metric, the fitted mean trend for healthy controls is shown as a gray solid line with shaded 95% confidence interval, and the fitted mean trend for AMD eyes at retinal locations without visible pathology (unremarkable regions) is shown as a red solid line with shaded 95% confidence interval. Lesion-associated measurements are overlaid as individual data points, color-coded by lesion type (drusen, subretinal drusenoid deposits (SDD), basal laminar deposits (BLamD), acquired vitelliform lesions (AVL), and geographic atrophy (GA)). Reference trends were estimated using control or unremarkable data only; lesion measurements were not included in model fitting and are shown for visualization. Because cohorts were age-matched, trends are displayed as functions of retinal eccentricity without explicit age stratification.

**Extended Data Table 1.** Linear mixed-effects model of RPE autofluorescence lifetime and cellular structural metrics in eyes of healthy subjects.

| Outcome | Age |  | Location |  | Gender | Race |
| --- | --- | --- | --- | --- | --- | --- |
| e | $\beta$ | p | $\beta$ | p | p | p |
| LSC $\tau_{m12}$ | 40.29 | <0.0001 | 23.44 | <0.0001 | 0.8407 | 0.2639 |
| LSC $\tau_1$ | 15.21 | <0.0001 | 14.26 | <0.0001 | 0.7114 | 0.7829 |
| LSC $\tau_2$ | 125.84 | 0.0038 | 64.05 | <0.0001 | 0.5486 | 0.2760 |
| LSC $\alpha_1$ | -1.12 | 0.2083 | -0.03 | 0.8641 | 0.3668 | 0.5865 |
| LSC $\alpha_2$ | 1.26 | 0.0422 | 0.11 | 0.5153 | 0.2623 | 0.8102 |
| SSC $\tau_{m12}$ | 29.52 | <0.0001 | 27.92 | <0.0001 | 0.5366 | 0.0919 |
| SSC $\tau_1$ | 15.64 | <0.0001 | 19.99 | <0.0001 | 0.4690 | 0.2672 |
| SSC $\tau_2$ | 160.27 | 0.0057 | 83.26 | <0.0001 | 0.6761 | 0.2332 |
| SSC $\alpha_1$ | 2.89 | 0.0528 | 1.03 | 0.0056 | 0.5708 | 0.5584 |
| SSC $\alpha_2$ | -2.48 | 0.0402 | -1.05 | 0.0052 | 0.6047 | 0.5185 |
| Cell density | -112.00 | 0.3304 | -495.01 | <0.0001 | 0.2137 | 0.1863 |
| Hexagonality | -0.02 | 0.0912 | -0.01 | 0.0782 | 0.2002 | 0.4319 |
| ICD | -0.01 | 0.9354 | 0.20 | <0.0001 | 0.0005 | 0.3325 |
| Neighbors | -0.02 | 0.3159 | -0.02 | 0.0043 | 0.8075 | 0.3273 |

Linear mixed-effects (LME) results from the full model including age, retinal eccentricity (location), gender, and race/ethnicity as fixed effects, with a subject-specific random intercept to account for repeated measurements within individuals. Outcomes included RPE autofluorescence lifetime parameters and cellular structural metrics, including cell density, hexagonal cell proportion (hexagonality), intercellular distance (ICD), and number of nearest neighbors (neighbors). Age and location were z-scored prior to model fitting; estimates therefore represent changes per standard deviation. For each outcome, standardized regression coefficients ( $\beta$ ) and two-sided p values are reported for age and retinal eccentricity. The overall effects of sex and race were assessed using likelihood ratio tests and are reported as p values. All models were fitted using maximum likelihood estimation. P values < 0.05 are highlighted in red. Full model outputs are provided in **Supplementary Tables 4–6**.

**Extended Data Table 2.** Core linear mixed-effects model of RPE autofluorescence lifetime and cell structure in healthy subjects.

| Outcome | N | Age |  |  |  |  | Location |  |  |  |  |
| --- | --- | --- | --- | --- | --- | --- | --- | --- | --- | --- | --- |
| | | $\beta$ | SE | p | CI- | CI+ | $\beta$ | SE | p | CI- | CI+ |
| LSC $\tau_{m12}$ | 152 | 43.0 | 4.3 | <0.0001 | 34.5 | 51.5 | 23.5 | 1.5 | <0.0001 | 20.5 | 26.4 |
| LSC $\tau_1$ | 152 | 14.8 | 1.9 | <0.0001 | 11.1 | 18.5 | 14.2 | 1.0 | <0.0001 | 12.2 | 16.3 |
| LSC $\tau_2$ | 152 | 132.7 | 30.3 | <0.0001 | 72.8 | 192.7 | 64.0 | 6.9 | <0.0001 | 50.5 | 77.6 |
| LSC $\alpha_1$ | 152 | -0.9 | 0.6 | 0.1440 | -2.1 | 0.3 | -0.04 | 0.2 | 0.8465 | -0.4 | 0.3 |
| LSC $\alpha_2$ | 152 | 1.3 | 0.4 | 0.0026 | 0.5 | 2.2 | 0.1 | 0.2 | 0.4859 | -0.2 | 0.5 |
| SSC $\tau_{m12}$ | 152 | 30.9 | 4.1 | <0.0001 | 22.8 | 39.1 | 28.0 | 1.9 | <0.0001 | 24.1 | 31.8 |
| SSC $\tau_1$ | 152 | 15.6 | 2.2 | <0.0001 | 11.4 | 19.9 | 20.0 | 1.4 | <0.0001 | 17.2 | 22.9 |
| SSC $\tau_2$ | 152 | 143.2 | 40.6 | 0.0006 | 62.9 | 223.5 | 83.3 | 11.0 | <0.0001 | 61.5 | 105.0 |
| SSC $\alpha_1$ | 152 | 2.2 | 1.0 | 0.0359 | 0.1 | 4.2 | 1.0 | 0.4 | 0.0059 | 0.3 | 1.7 |
| SSC $\alpha_2$ | 152 | -1.7 | 0.8 | 0.0487 | -3.3 | 0.0 | -1.0 | 0.4 | 0.0056 | -1.8 | -0.3 |
| Cell density | 152 | -221.2 | 84.2 | 0.0095 | -387.6 | -54.8 | -493.0 | 55.6 | <0.0001 | -602.9 | -383.1 |
| Hexagonality | 152 | -0.02 | 0.01 | 0.0140 | -0.04 | 0.00 | -0.01 | 0.01 | 0.1065 | -0.03 | 0.00 |
| ICD | 152 | 0.1 | 0.1 | 0.3240 | -0.1 | 0.2 | 0.2 | 0.0 | <0.0001 | 0.1 | 0.3 |
| Neighbors | 152 | -0.01 | 0.01 | 0.6021 | -0.03 | 0.02 | -0.02 | 0.01 | 0.0041 | -0.03 | 0.01 |

**Extended Data Table 3.** AOFLIO measurements of RPE autofluorescence lifetime in eyes with age-related macular degeneration compared with age-matched healthy controls.

| Outcome | Unremarkable |  | SDD |  | Drusen |  | AVL |  | BlamD |  |
| --- | --- | --- | --- | --- | --- | --- | --- | --- | --- | --- |
| | $\beta$ | p | $\beta$ | p | $\beta$ | p | $\beta$ | p | $\beta$ | p |
| LSC $\tau_{m12}$ | 72.96 | 0.0009 | 81.81 | 0.0003 | 87.38 | <0.0001 | 35.91 | 0.1542 | 77.01 | 0.0012 |
| LSC $\tau_1$ | 34.60 | 0.0005 | 31.51 | 0.0024 | 34.70 | 0.0005 | -7.94 | 0.5136 | 35.17 | 0.0016 |
| LSC $\tau_2$ | 119.11 | 0.0202 | 157.44 | 0.0035 | 124.89 | 0.0162 | 131.11 | 0.0355 | 84.79 | 0.1385 |
| LSC $\alpha_1$ | -3.06 | 0.0304 | -4.28 | 0.0032 | -4.70 | 0.0010 | -3.36 | 0.0325 | -4.70 | 0.0018 |
| LSC $\alpha_2$ | 2.63 | 0.0401 | 3.45 | 0.0086 | 3.95 | 0.0023 | 1.97 | 0.1646 | 3.77 | 0.0056 |
| SSC $\tau_{m12}$ | 39.45 | 0.0308 | 62.80 | 0.0012 | 58.46 | 0.0017 | 15.13 | 0.5052 | 49.77 | 0.0166 |
| SSC $\tau_1$ | 24.65 | 0.0169 | 27.74 | 0.0106 | 25.40 | 0.0152 | -22.13 | 0.0793 | 24.46 | 0.0347 |
| SSC $\tau_2$ | 53.00 | 0.5402 | 220.02 | 0.0155 | 121.23 | 0.1667 | 178.40 | 0.0871 | 84.89 | 0.3774 |
| SSC $\alpha_1$ | -2.59 | 0.1409 | -2.10 | 0.2494 | -3.73 | 0.0361 | -3.16 | 0.1247 | -3.06 | 0.1114 |
| SSC $\alpha_2$ | 2.21 | 0.0855 | 1.38 | 0.3122 | 3.46 | 0.0084 | 2.20 | 0.1788 | 3.01 | 0.0420 |
| Cell density | -604.73 | 0.0033 | -943.67 | <0.0001 | -638.92 | 0.0024 | -318.15 | 0.2477 | -352.25 | 0.1483 |
| Hexagonality | -0.005 | 0.8525 | 0.01 | 0.6552 | 0.01 | 0.7553 | -0.06 | 0.1555 | 0.05 | 0.1415 |
| ICD | 1.48 | <0.0001 | 2.12 | <0.0001 | 1.47 | <0.0001 | 1.30 | 0.0003 | 1.19 | 0.0002 |
| Neighbors | 0.05 | 0.0273 | 0.00 | 0.8886 | 0.02 | 0.5130 | 0.01 | 0.8173 | 0.06 | 0.0409 |

Group-level differences in RPE autofluorescence lifetime parameters ( $\tau_{m12}$ , $\tau_1$ , $\tau_2$ , $\alpha_1$ , $\alpha_2$ in both channels) and cellular structural metrics (cell density, hexagonal cell proportion (Hexagonality), intercellular distance (ICD), and number of nearest neighbors (Neighbors)) were evaluated using linear mixed-effects models with retinal eccentricity included as a fixed-effect covariate and a subject-specific random intercept to account for within-eye correlation. In this table, healthy control regions serve as the reference group, and fixed-effect estimates ( $\beta$ ) represent the estimated difference between each AMD region (non-lesion or lesion-associated regions including SDD, drusen, AVL, and BlamD) and healthy controls after adjusting for eccentricity. Positive $\beta$ values indicate higher outcome values relative to healthy controls, whereas negative $\beta$ values indicate lower values. Statistical inference was performed at the group level using all available measurement locations. Lesions contributing a single measurement location were included as individual observations but were not interpreted as providing lesion-level statistical evidence. Two-sided p-values are reported without adjustment for multiple comparisons, and values with $p < 0.05$ are highlighted in red. Complete pairwise contrast results are provided in **Supplementary Tables 10–12**.

**Extended Data Table 4.** AOFLIO measurements of RPE autofluorescence lifetime in regions with age-related macular degeneration lesions compared with unremarkable regions.

| Outcome | SDD |  | Drusen |  | AVL |  | BlamD |  |
| --- | --- | --- | --- | --- | --- | --- | --- | --- |
| | $\beta$ | p | $\beta$ | p | $\beta$ | p | $\beta$ | p |
| LSC $\tau_{m12}$ | 8.85 | 0.3462 | 14.42 | 0.0730 | -37.04 | 0.0110 | 4.05 | 0.7219 |
| LSC $\tau_1$ | -3.09 | 0.5642 | 0.10 | 0.9828 | -42.54 | <0.0001 | 0.57 | 0.9300 |
| LSC $\tau_2$ | 38.32 | 0.1456 | 5.77 | 0.7964 | 11.99 | 0.7673 | -34.33 | 0.2811 |
| LSC $\alpha_1$ | -1.22 | 0.0169 | -1.64 | 0.0002 | -0.30 | 0.7021 | -1.64 | 0.0082 |
| LSC $\alpha_2$ | 0.82 | 0.0707 | 1.32 | 0.0008 | -0.66 | 0.3494 | 1.15 | 0.0386 |
| SSC $\tau_{m12}$ | 23.35 | 0.0212 | 19.01 | 0.0271 | -24.31 | 0.1185 | 10.32 | 0.3975 |
| SSC $\tau_1$ | 3.09 | 0.5672 | 0.75 | 0.8704 | -46.79 | <0.0001 | -0.19 | 0.9766 |
| SSC $\tau_2$ | 167.02 | <0.0001 | 68.23 | 0.0625 | 125.40 | 0.0586 | 31.89 | 0.5390 |
| SSC $\alpha_1$ | 0.49 | 0.5349 | -1.14 | 0.0908 | -0.57 | 0.6409 | -0.47 | 0.6222 |
| SSC $\alpha_2$ | -0.83 | 0.2684 | 1.24 | 0.0515 | -0.01 | 0.9914 | 0.80 | 0.3786 |
| Cell density | -338.94 | 0.0137 | -34.19 | 0.7665 | 286.57 | 0.1737 | 252.48 | 0.1259 |
| Hexagonality | 0.02 | 0.4835 | 0.01 | 0.5185 | -0.05 | 0.1489 | 0.05 | 0.0636 |
| ICD | 0.64 | 0.0003 | -0.01 | 0.9554 | -0.18 | 0.5119 | -0.29 | 0.1785 |
| Neighbors | -0.05 | 0.0417 | -0.04 | 0.0462 | -0.04 | 0.2054 | 0.01 | 0.6547 |

Group-level differences in RPE autofluorescence lifetime parameters ( $\tau_{m12}$ , $\tau_1$ , $\tau_2$ , $\alpha_1$ , $\alpha_2$ in both channels) and cellular structural metrics (RPE cell density, hexagonal cell proportion (Hexagonality), intercellular distance (ICD), and number of nearest neighbors (Neighbors)) were evaluated using linear mixed-effects models with retinal eccentricity included as a fixed-effect covariate and a subject-specific random intercept to account for within-eye correlation. In this table, AMD non-lesion (unremarkable) regions serve as the reference group, and fixed-effect estimates ( $\beta$ ) represent the estimated difference between each AMD region (non-lesion or lesion-associated regions including SDD, drusen, AVL, and BlamD) and healthy controls after adjustment for eccentricity. Positive $\beta$ values indicate higher outcome values relative to healthy controls, whereas negative $\beta$ values indicate lower values. Statistical inference was performed at the group level using all available measurement locations. Lesions contributing a single measurement location were included as individual observations but were not interpreted as providing lesion-level statistical evidence. Two-sided p-values are reported without adjustment for multiple comparisons, and values with $p < 0.05$ are highlighted in red. Complete pairwise contrast results are provided in **Supplementary Tables 10–12**.

## Supplementary Materials

### Supplementary note 1: Human subjects, and Standard Clinical Characterization

**1) Human subjects**

Study patients with AMD and age-similar subjects with normal retinas were recruited from the clinical research registry of Doheny Eye Center, University of California – Los Angeles. Inclusion criteria: the patients have been diagnosed with AMD previously. Exclusion criteria included diabetes, history of retinal vascular occlusions, and any signs or history of hereditary retinal dystrophy. Subjects were also excluded for reasons that might potentially prevent successful imaging, such as poor fixation, significant media opacity, irregular pupil shape, poor dilation, or refractive errors beyond ±8 D spherical and ±3 D cylinder. The inclusion criteria for normal comparison subjects were the same, with the additional criteria of age greater than 50 years, no clinically significant cataract, and best-corrected visual acuity (BCVA) of 20/40 or better.

**Supplementary Tables 1–3** summarize cohort demographics, with no significant age difference between the AMD and control groups (Welch’s t-test p = 0.1021).

**Supplementary Table 1.** Health subject characteristics.

| Race/Ethnicity | Gender |  | Total | Age (years) |
| --- | --- | --- | --- | --- |
|  | Females | Males |  |  |
| White non-Hispanic | 7 | 5 | 12 | 67.58 ± 10.19 |
| Hispanic | 6 | 3 | 9 | 39.67 ± 11.74 |
| Asian | 8 | 7 | 15 | 42.87 ± 11.46 |
| **Total** | 21 | 15 | 36 | $p < 0.0001$ |
| **Age (years)** | 50.57 ± 10.90 | 49.93 ± 12.68 | $p = 0.9101$ | |

**Supplementary Table 2.** AMD subject characteristics.

| Race/Ethnicity | Gender |  | Total | Age (years) |
| --- | --- | --- | --- | --- |
|  | Females | Males |  |  |
| White non-Hispanic | 6 | 7 | 13 | 76.62 ± 7.18 |
| **Age (years)** | 74.5 ± 7.48 | 78.43 ± 6.43 | $p = 0.3376$ | |

**Supplementary Table 3.** Health control group character.

| Race/Ethnicity | Gender |  | Total | Age (years) |
| --- | --- | --- | --- | --- |
|  | Females | Males |  |  |
| White non-Hispanic | 6 | 3 | 9 | 71.89 ± 5.90 |
| **Age (years)** | 71.67 ± 8.59 | 72.33 ± 8.39 | $p = 0.9082$ | |

**2) Clinical multimodal imaging**

The enrolled subjects underwent multimodal ophthalmic imaging, including color fundus photography, fundus autofluorescence imaging (excitation, 488 nm; emission > 600 nm) using an ultra-widefield (UWF™) retinal imager (California, Optos, Inc., Marlborough, MA). Retinal cross-sections were imaged with the Spectralis SD-OCT (λ = 870 nm; acquisition speed, 40,000 A-scans per seconds; scan depth, 1.9 mm; digital depth resolution, 3.5 µm per pixel in tissue; lateral resolution in tissue 14 µm). In each study eye, 97 B-scans were acquired across a 20° X 20° area of the central macula to create a volume. Prior to imaging, subjects’ pupils were dilated, and accommodation was paralyzed by instillation of topical tropicamide (1%) and phenylephrine hydrochloride (2.5%) to ensure optimal imaging conditions. A retinal specialist reviewed the images to confirm the retinal health status. Axial length measurements were obtained using an ocular biometer (IOL Master; Carl Zeiss Meditec, Germany).

**3) AOSLO Image processing**

Adaptive optics scanning laser ophthalmoscopy (AOSLO) images were corrected for distortions caused by resonant scanner nonlinearities and eye motion using custom software. Registered frames were averaged to improve signal-to-noise ratio. Images acquired at different retinal locations were manually aligned on a cell-to-cell basis to generate continuous AOSLO montages (Adobe Photoshop, Adobe Systems Inc., Mountain View, CA). The retinal pixel scale of AOSLO images was calibrated using images of a precision dot grid placed at the retinal plane of a model eye. The spatial extent of retinal regions affected by AMD lesions was measured at multiple locations using the Ruler tool in Photoshop and averaged across measurements.

**4) Multimodal image registration**

Color fundus photographs and infrared (IR), red-free (RF), and autofluorescence images acquired with the Spectralis scanning laser ophthalmoscope were manually registered using retinal vessels and capillaries as invariant landmarks. These images were subsequently magnified and registered to the AOSLO montages using the same vascular landmarks. Lesions identified in clinical imaging were thus localized within the corresponding high-resolution AOSLO images and examined at cellular resolution.

**5) AMD lesion characterization**

AMD lesion types and clinically unremarkable retinal regions were defined based on combined evaluation of en face fundus imaging and SD-OCT, following established clinical criteria.

Representative examples of lesion localization across imaging modalities are shown in **Supplementary Figs. 3–6**.

**Drusen**: Drusen were identified as extracellular deposits located between the retinal pigment epithelium (RPE) and Bruch’s membrane. On color fundus photography, drusen appeared as discrete yellow-white lesions. On IR reflectance imaging, they appeared as hyperreflective or hyporeflective spots, and on FAF as variable autofluorescent signals. On SD-OCT, drusen were characterized by elevations of the RPE band with underlying hyperreflective material.

**SDDs**: SDDs were identified based on their presence in at least two en face imaging modalities and confirmation on SD-OCT. En face imaging revealed SDD as an interlacing ribbon-like pattern or clusters of dot-like lesions, appearing yellow white on color fundus photography, hyporeflective or hyperreflective on IR reflectance, and hypoautofluorescent on FAF. On SD-OCT, SDD appeared as hyperreflective mounds located internal to the RPE. Axial microstructure and localization were evaluated using the CAM nomenclature.^96^

**AVL**: AVL were identified as focal accumulations of hyperautofluorescent material at or above the level of the RPE. On color fundus photography, AVL appeared as yellowish subretinal lesions. On FAF, they exhibited marked hyperautofluorescence, and on SD-OCT, they appeared as hyperreflective subretinal material overlying the RPE.

**BLamD:** BLamD were identified based on characteristic SD-OCT features, including thickening or separation of the RPE–Bruch’s membrane complex. En face imaging features were variable, but BLamD were distinguished from drusen and SDD based on their laminar location and OCT morphology.

**GA**: GA was identified by sharply demarcated regions of RPE and outer retinal atrophy. On color fundus photography, GA appeared as well-defined areas of depigmentation with visible underlying choroidal vessels. FAF imaging showed hypoautofluorescent regions corresponding to RPE loss, and SD-OCT revealed loss of the RPE band and overlying photoreceptor layers. GA regions were included for representative imaging analyses but contributed limited quantitative measurements.

**Clinically unremarkable regions**: Unremarkable regions were defined as retinal areas within AMD eyes that showed no visible pathological features on color fundus photography, FAF, IR reflectance imaging, or OCT.

### Supplementary note 2. Correcting for ocular chromatic aberration

The eye’s chromatic aberration makes AOSLO and AOFLIO imaging lights focus on different longitudinal and transverse points of the retina. The longitudinal chromatic aberration (LCA) and transverse chromatic aberration (TCA) were minimized by the “dual source-retina-detector conjugate” strategy^68–71^ First, we adjusted the NIR AOSLO (S_λ2_, λ = 790 nm) to focus on the cone photoreceptor inner segment layer, showing a clear cone mosaic. Then we used the PMT 7422A-40 to collect the reflectance of the excitation light (S_λ1_, λ = 473 nm) to produce an image of the cone mosaic. Next, we adjusted the light source and detector of the AOSLO (λ = 790 nm) to align its image with the PMT 7422A-40 image.

**Supplementary Fig. 1.**
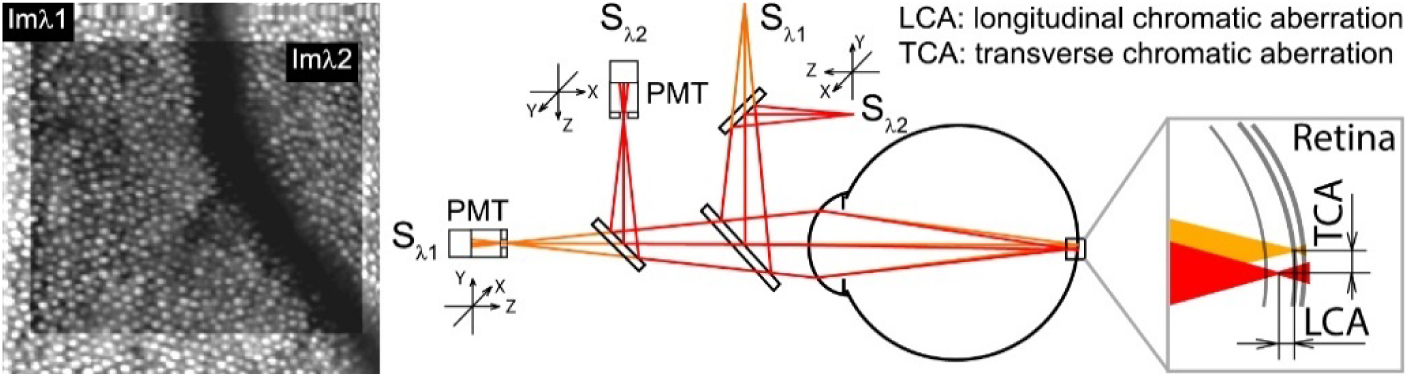
Dual source-retina-detector conjugate” strategy for minimizing ocular chromatic aberration.

### Supplementary note 3. Post-processing of AOFLIO data for autofluorescence photon intensity and lifetime imaging

**Supplementary Fig. 2.**
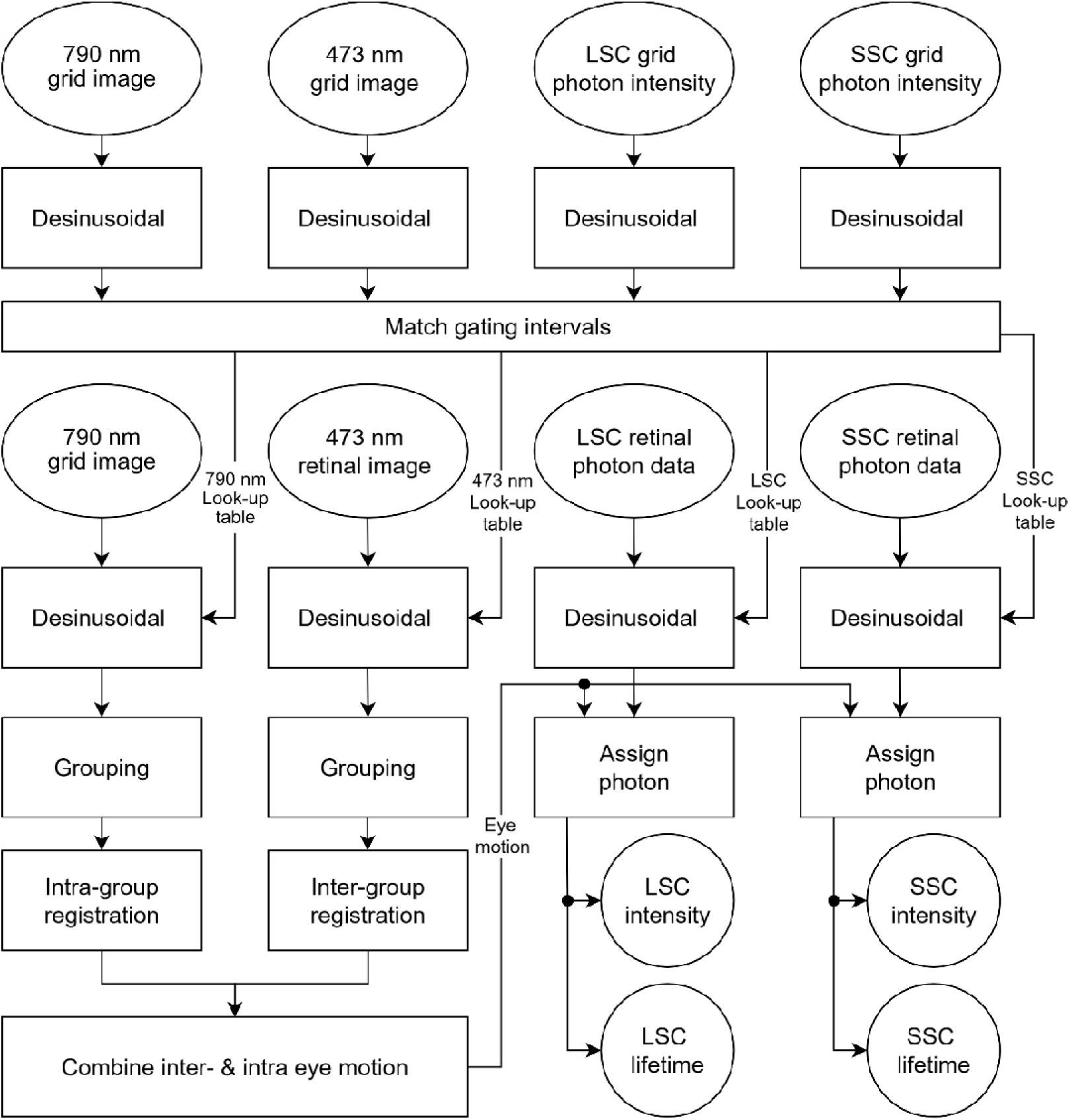
Post-processing of AOFLIO data for autofluorescence photon intensity and lifetime imaging. The simultaneously acquired four-channel images (AOSLO reflectance at 790 nm and 473 nm, AOFLIO fluorescence LSC and SSC channels) each utilize standard grid images as references to correct sinusoidal scanning distortion. Reflectance image frames are grouped and registered to measure eye motion. Photon events are then precisely assigned using sinusoidal distortion correction lookup tables and eye motion data, producing accurate photon intensity and fluorescence lifetime images.

### Supplementary note 4. Standard multimodal imaging of eyes with AMD

**Supplementary Fig. 3.**
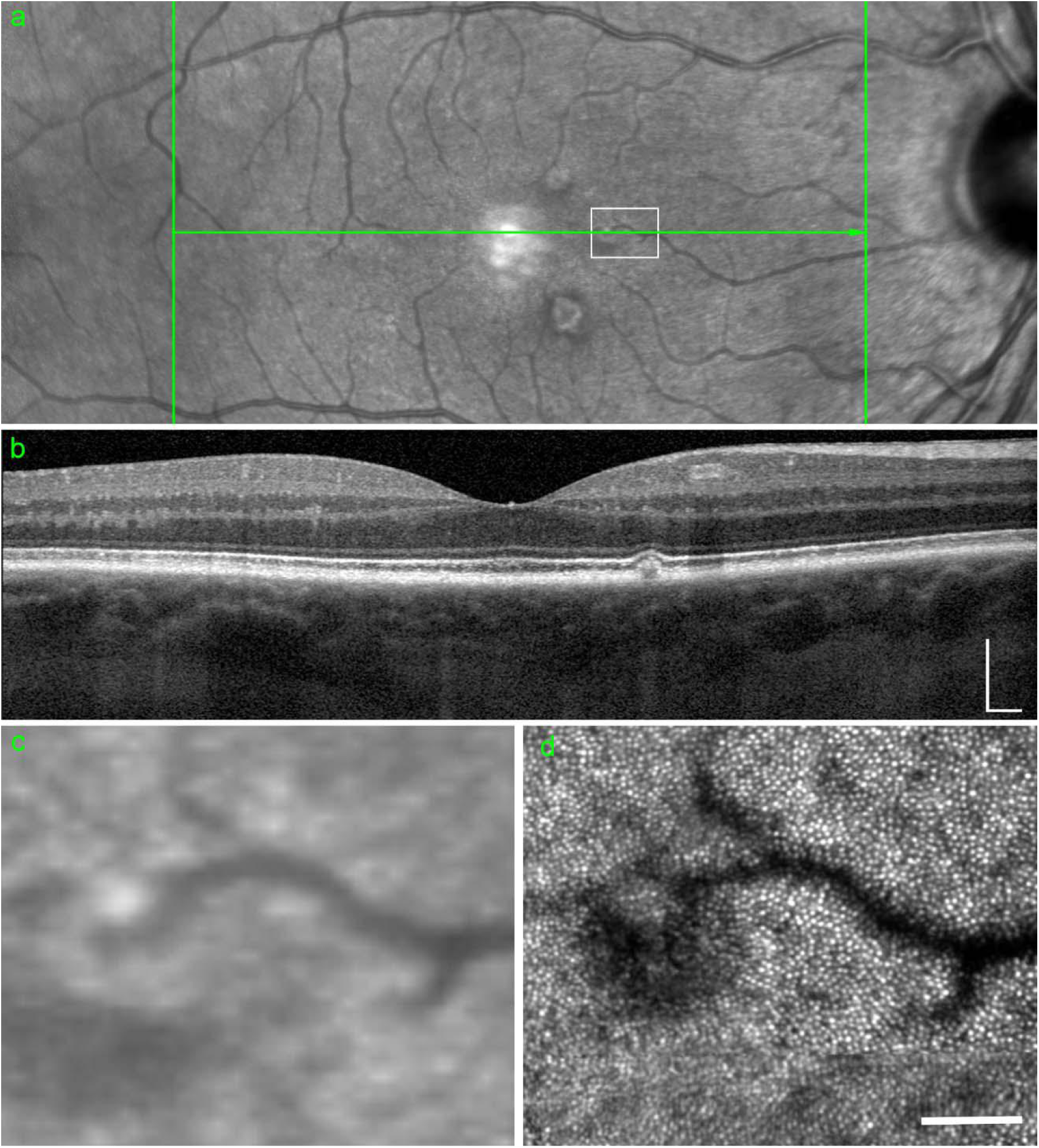
Clinical fundus, OCT, and AOSLO images of drusen. (a) Fundus photograph. The green line indicates the location of the OCT B-scan, and the white box marks the region containing drusen lesions. (b) Corresponding OCT B-scan image. Scale bar, 200 µm (c,d) Magnified view of the boxed region in (a) and the corresponding 790-nm AOSLO reflectance image of the same field of view. Scale bar, 100 µm.

**Supplementary Fig. 4.**
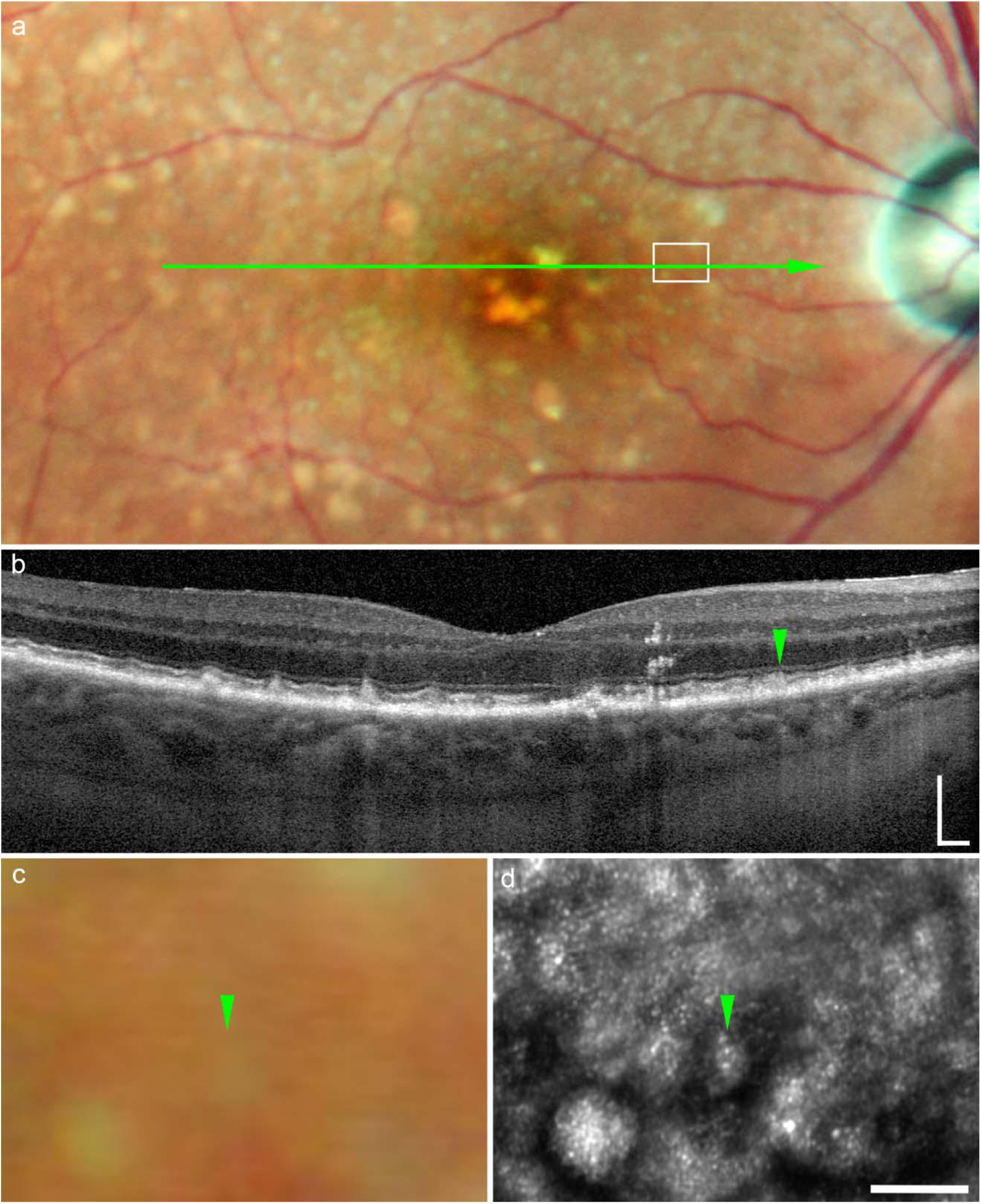
Clinical fundus, OCT, and AOSLO images of subretinal drusenoid deposits (SDD). (a) Color fundus photograph. The green line indicates the OCT B-scan location, and the white box marks the region containing SDD lesions. (b) Corresponding OCT B-scan image. Scale bar, 200 µm. (c,d) Magnified view of the boxed region in (a) and the corresponding 790-nm AOSLO reflectance image of the same field of view. Scale bar, 100 µm. Green arrows indicate the SDD lesion shown in Fig. 5.

**Supplementary Fig. 5.**
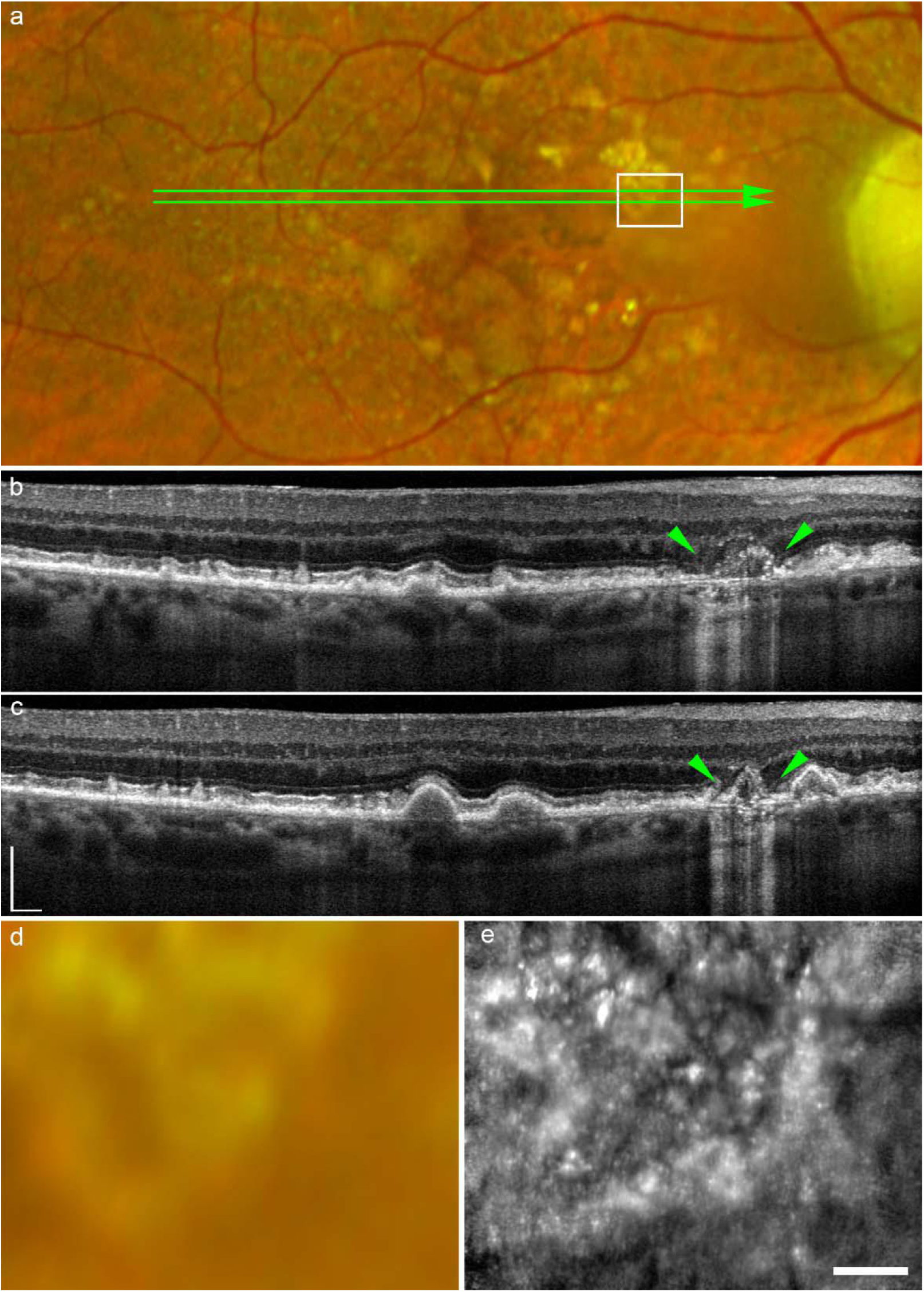
Clinical fundus, OCT, and AOSLO images of collapsing drusen. (a) Color fundus photograph. Two green lines indicate the OCT B-scan locations corresponding to (b) and (c). The white box marks the region containing collapsing drusen. (b,c) Corresponding OCT B-scan images. Green arrows indicate the boundary regions of collapsing drusen. Scale bar, 200 µm. (d,e) Magnified view of the boxed region in (a) and the corresponding 790-nm AOSLO reflectance image of the same field of view. Scale bar, 100 µm.

**Supplementary Fig. 6.**
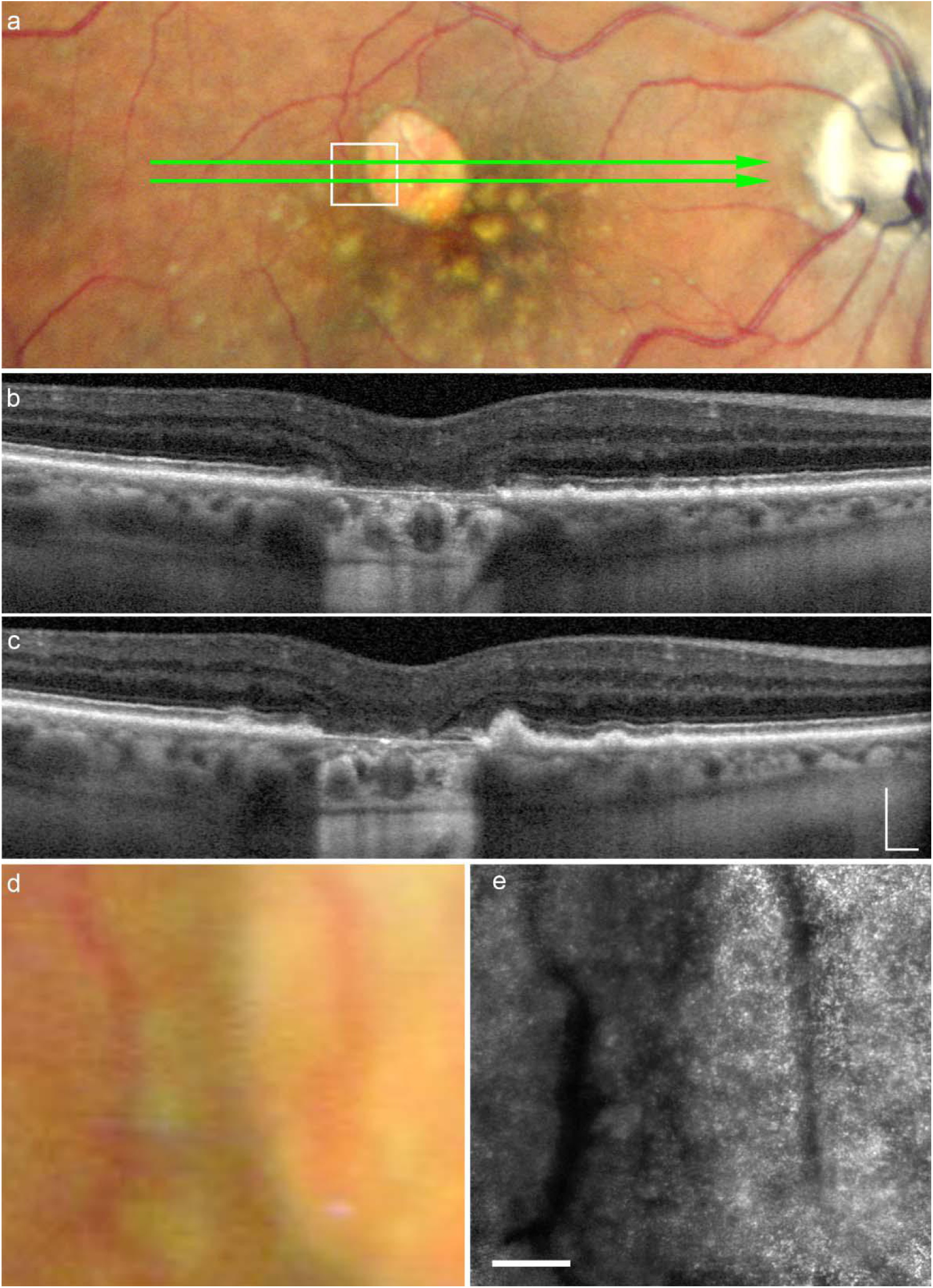
Clinical fundus, OCT, and AOSLO images of geographic atrophy (GA). (a) Color fundus photograph. Two green lines indicate the OCT B-scan locations corresponding to (b) and (c). The white box marks the region containing GA lesions. (b,c) Corresponding OCT B-scan images. Green arrows indicate the boundary regions of GA. Scale bar, 200 µm. (c,d) Magnified view of the boxed region in (a) and the corresponding 790-nm AOSLO reflectance image of the same field of view. Scale bar, 100 µm.

### Supplementary note 5. Linear mixed-effects modeling for normative reference construction in healthy eyes

To establish normative reference in healthy eyes, linear mixed-effects (LME) models were fitted separately for three outcome domains: LSC fluorescence lifetime parameters (**Supplementary Table 4**), SSC fluorescence lifetime parameters (**Supplementary Table 5**), and cellular structural metrics (**Supplementary Table 6**). Across all models, age and retinal eccentricity were included as continuous fixed effects, sex and race as categorical fixed effects, and a subject-specific random intercept was used to account for repeated measurements within individuals. Models were fitted using maximum likelihood estimation. Reported statistics include regression coefficients (β), two-sided p values, 95% confidence intervals, variance inflation factors (VIF), marginal and conditional R², and intraclass correlation coefficients (ICC). Age and retinal eccentricity were mean centered prior to analysis. Sex and race were coded relative to reference categories defined in the Methods.

**Supplementary Table 4.** Full linear mixed-effects model outputs for LSC fluorescence lifetime parameters.

| Outcome | Term | $\beta$ | p Value | CI- | CI+ | VIF | $R^2_m$ | $R^2_c$ | ICC |
| --- | --- | --- | --- | --- | --- | --- | --- | --- | --- |
| LSC $\tau_{m12}$ | (Intercept) | 265.50 | <0.0001 | 246.24 | 284.75 | | | | |
|  | Age_c | 40.29 | <0.0001 | 28.25 | 52.34 | 2.04 |  |  |  |
|  | Loc_c | 23.44 | <0.0001 | 20.50 | 26.38 | 1.03 |  |  |  |
|  | Sex_2 | -1.84 | 0.8407 | -19.91 | 16.23 | 0.54 | 0.7575 | 0.9008 | 0.5901 |
|  | Race_2 | 14.83 | 0.1501 | -5.43 | 35.10 | 1.04 |  |  |  |
|  | Race_3 | 15.42 | 0.2574 | -11.38 | 42.22 | 1.71 |  |  |  |
| LSC $\tau_1$ | (Intercept) | 188.58 | <0.0001 | 179.86 | 197.30 | | | | |
|  | Age_c | 15.21 | <0.0001 | 9.75 | 20.67 | 2.04 |  |  |  |
|  | Loc_c | 14.26 | <0.0001 | 12.19 | 16.33 | 1.03 |  |  |  |
|  | Sex_2 | -1.54 | 0.7114 | -9.74 | 6.66 | 0.54 | 0.6302 | 0.7482 | 0.3191 |
|  | Race_2 | 3.16 | 0.4945 | -5.96 | 12.29 | 1.04 |  |  |  |
|  | Race_3 | 0.29 | 0.9619 | -11.88 | 12.47 | 1.71 |  |  |  |
| LSC $\tau_2$ | (Intercept) | 714.74 | <0.0001 | 579.61 | 849.87 | | | | |
|  | Age_c | 125.84 | 0.0038 | 41.30 | 210.38 | 2.04 |  |  |  |
|  | Loc_c | 64.05 | <0.0001 | 50.50 | 77.61 | 1.03 |  |  |  |
|  | Sex_2 | -38.54 | 0.5486 | -165.23 | 88.15 | 0.54 | 0.4442 | 0.8804 | 0.7849 |
|  | Race_2 | 112.64 | 0.1205 | -29.88 | 255.17 | 1.04 |  |  |  |
|  | Race_3 | 76.67 | 0.4212 | -111.20 | 264.54 | 1.71 |  |  |  |
| LSC $\alpha_1$ | (Intercept) | 82.40 | <0.0001 | 79.60 | 85.20 | | | | |
|  | Age_c | -1.12 | 0.2083 | -2.87 | 0.63 | 2.04 |  |  |  |
|  | Loc_c | -0.03 | 0.8641 | -0.40 | 0.34 | 1.03 |  |  |  |
|  | Sex_2 | -1.20 | 0.3668 | -3.83 | 1.42 | 0.54 | 0.0937 | 0.6967 | 0.6653 |
|  | Race_2 | 1.41 | 0.3456 | -1.54 | 4.36 | 1.04 |  |  |  |
|  | Race_3 | 1.30 | 0.5098 | -2.59 | 5.20 | 1.71 |  |  |  |
| LSC $\alpha_2$ | (Intercept) | 14.87 | <0.0001 | 12.92 | 16.81 | | | | |
|  | Age_c | 1.26 | 0.0422 | 0.05 | 2.48 | 2.04 |  |  |  |
|  | Loc_c | 0.11 | 0.5153 | -0.22 | 0.44 | 1.03 |  |  |  |
|  | Sex_2 | 1.04 | 0.2623 | -0.79 | 2.87 | 0.54 | 0.1975 | 0.6175 | 0.5234 |
|  | Race_2 | -0.67 | 0.5189 | -2.72 | 1.38 | 1.04 |  |  |  |
|  | Race_3 | -0.16 | 0.9059 | -2.87 | 2.55 | 1.71 |  |  |  |

P value less than 0.05 highlighted in red.

**Supplementary Table 5.** Full linear mixed-effects model outputs for SSC fluorescence lifetime parameters.

| Outcome | Term | $\beta$ | p Value | CI- | CI+ | VIF | R<sup>2</sup>m | R<sup>2</sup>c | ICC |
| --- | --- | --- | --- | --- | --- | --- | --- | --- | --- |
| SSC $\tau_{m12}$ | (Intercept) | 204.28 | <0.0001 | 186.50 | 222.05 | | | | |
|  | Age_c | 29.52 | <0.0001 | 18.40 | 40.65 | 2.04 |  |  |  |
|  | Loc_c | 27.92 | <0.0001 | 24.11 | 31.73 | 1.03 |  |  |  |
|  | Sex_2 | -5.24 | 0.5366 | -21.94 | 11.47 | 0.54 | 0.6652 | 0.7941 | 0.3849 |
|  | Race_2 | 20.09 | 0.0348 | 1.45 | 38.72 | 1.04 |  |  |  |
|  | Race_3 | 14.07 | 0.2639 | -10.72 | 38.86 | 1.71 |  |  |  |
| SSC $\tau_1$ | (Intercept) | 153.17 | <0.0001 | 143.71 | 162.63 | | | | |
|  | Age_c | 15.64 | <0.0001 | 9.70 | 21.57 | 2.04 |  |  |  |
|  | Loc_c | 19.99 | <0.0001 | 17.15 | 22.82 | 1.03 |  |  |  |
|  | Sex_2 | -3.28 | 0.4690 | -12.20 | 5.65 | 0.54 | 0.6246 | 0.6914 | 0.1780 |
|  | Race_2 | 8.11 | 0.1059 | -1.74 | 17.97 | 1.04 |  |  |  |
|  | Race_3 | 3.83 | 0.5688 | -9.43 | 17.09 | 1.71 |  |  |  |
| SSC $\tau_2$ | (Intercept) | 540.53 | <0.0001 | 359.94 | 721.13 | | | | |
|  | Age_c | 160.27 | 0.0057 | 47.29 | 273.24 | 2.04 |  |  |  |
|  | Loc_c | 83.26 | <0.0001 | 61.54 | 104.98 | 1.03 |  |  |  |
|  | Sex_2 | -35.87 | 0.6761 | -205.24 | 133.50 | 0.54 | 0.3554 | 0.8143 | 0.7119 |
|  | Race_2 | 163.89 | 0.0909 | -26.44 | 354.23 | 1.04 |  |  |  |
|  | Race_3 | 32.95 | 0.7958 | -218.23 | 284.12 | 1.71 |  |  |  |
| SSC $\alpha_1$ | (Intercept) | 82.05 | <0.0001 | 77.36 | 86.73 | | | | |
|  | Age_c | 2.89 | 0.0528 | -0.04 | 5.83 | 2.04 |  |  |  |
|  | Loc_c | 1.03 | 0.0056 | 0.31 | 1.75 | 1.03 |  |  |  |
|  | Sex_2 | -1.26 | 0.5708 | -5.66 | 3.13 | 0.54 | 0.1390 | 0.6735 | 0.5860 |
|  | Race_2 | 2.29 | 0.3606 | -2.64 | 7.22 | 1.04 |  |  |  |
|  | Race_3 | -1.00 | 0.7633 | -7.52 | 5.52 | 1.71 |  |  |  |
| SSC $\alpha_2$ | (Intercept) | 14.90 | <0.0001 | 11.12 | 18.69 | | | | |
|  | Age_c | -2.48 | 0.0402 | -4.85 | -0.11 | 2.04 |  |  |  |
|  | Loc_c | -1.05 | 0.0052 | -1.79 | -0.32 | 1.03 |  |  |  |
|  | Sex_2 | 0.93 | 0.6047 | -2.62 | 4.49 | 0.54 | 0.1179 | 0.5147 | 0.4497 |
|  | Race_2 | -1.49 | 0.4603 | -5.47 | 2.49 | 1.04 |  |  |  |
|  | Race_3 | 1.72 | 0.5215 | -3.56 | 6.99 | 1.71 |  |  |  |

P value less than 0.05 highlighted in red.

**Supplementary Table 6.**
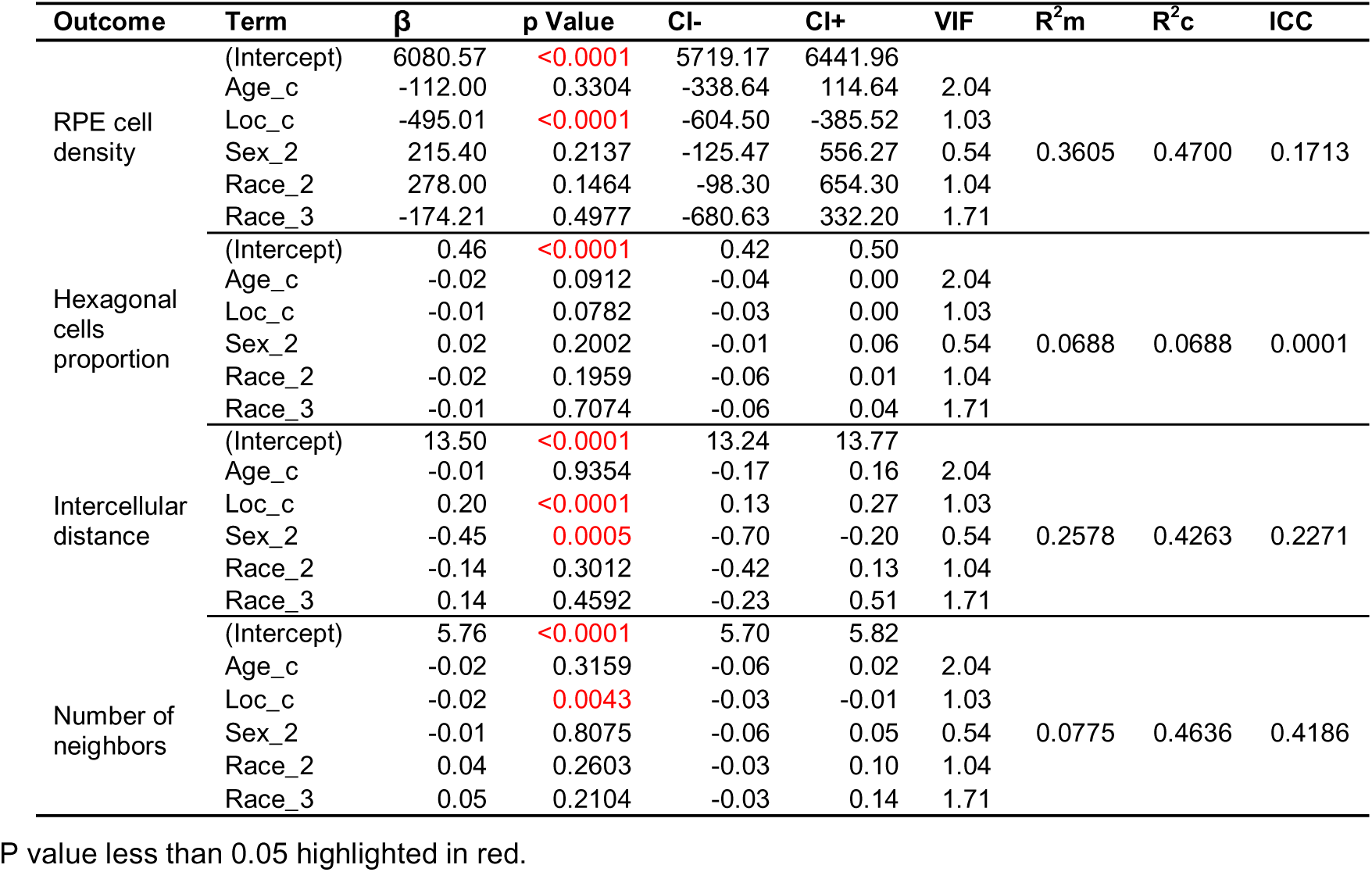
Full linear mixed-effects model outputs for cellular structural metrics.

### Supplementary note 6. Term-removal–based model selection for normative reference construction

To identify parsimonious core models for normative reference construction, likelihood ratio test (LRT)–based term removal analyses were performed for each outcome. Starting from the full linear mixed-effects (LME) models described in Supplementary Note S4, fixed-effect terms (age, retinal eccentricity/location, sex, race, and the combined sex + race term) were sequentially removed while holding the random-effects structure constant. For each reduced model, likelihood ratio statistics, changes in information criteria (ΔAIC and ΔBIC), and false discovery rate (FDR)–adjusted p values (Benjamini–Hochberg) were computed. Term retention decisions were based on whether removal of the term significantly worsened model fit (FDR-adjusted p < 0.05). Terms whose removal did not degrade model fit were considered nonessential and excluded from the core normative reference models. Results are reported separately for LSC fluorescence lifetime parameters (**Supplementary Table 7**), SSC fluorescence lifetime parameters (**Supplementary Table 8**), and cellular structural metrics (**Supplementary Table 9**).

**Supplementary Table 7.** Term-removal model comparison for LSC fluorescence lifetime parameters.

| Outcome | Term dropped | LRT p | FDR-adjusted p | LR statistic | ΔAIC | ΔBIC | Keep |
| --- | --- | --- | --- | --- | --- | --- | --- |
| LSC $\tau_{m12}$ | Age | 0.0000 | 0.0000 | 25.93 | 23.93 | 20.91 | Yes |
|  | Location | 0.0000 | 0.0000 | 136.24 | 134.24 | 131.21 | Yes |
|  | Sex | 0.8404 | 0.8404 | 0.04 | -1.96 | -4.98 | No |
|  | Race | 0.2761 | 0.6054 | 2.57 | -1.43 | -7.47 | No |
|  | Sex+Race | 0.4566 | 0.7645 | 2.60 | -3.40 | -12.47 | No |
| LSC $\tau_1$ | Age | 0.0000 | 0.0000 | 20.22 | 18.22 | 15.19 | Yes |
|  | Location | 0.0000 | 0.0000 | 117.23 | 115.23 | 112.20 | Yes |
|  | Sex | 0.7110 | 0.8295 | 0.14 | -1.86 | -4.89 | No |
|  | Race | 0.7849 | 0.8109 | 0.48 | -3.52 | -9.56 | No |
|  | Sex+Race | 0.8983 | 0.8983 | 0.59 | -5.41 | -14.48 | No |
| LSC $\tau_2$ | Age | 0.0062 | 0.0172 | 7.50 | 5.50 | 2.48 | Yes |
|  | Location | 0.0000 | 0.0000 | 65.78 | 63.78 | 60.75 | Yes |
|  | Sex | 0.5489 | 0.8295 | 0.36 | -1.64 | -4.66 | No |
|  | Race | 0.2883 | 0.6054 | 2.49 | -1.51 | -7.56 | No |
|  | Sex+Race | 0.4290 | 0.7645 | 2.77 | -3.23 | -12.30 | No |
| LSC $\alpha_1$ | Age | 0.2126 | 0.2705 | 1.55 | -0.45 | -3.47 | No |
|  | Location | 0.8639 | 0.8639 | 0.03 | -1.97 | -4.99 | No |
|  | Sex | 0.3685 | 0.8295 | 0.81 | -1.19 | -4.22 | No |
|  | Race | 0.5906 | 0.6890 | 1.05 | -2.95 | -8.99 | No |
|  | Sex+Race | 0.6137 | 0.7645 | 1.81 | -4.19 | -13.27 | No |
| LSC $\alpha_2$ | Age | 0.0476 | 0.0833 | 3.92 | 1.92 | -1.10 | No |
|  | Location | 0.5146 | 0.5542 | 0.42 | -1.58 | -4.60 | No |
|  | Sex | 0.2652 | 0.8295 | 1.24 | -0.76 | -3.78 | No |
|  | Race | 0.8109 | 0.8109 | 0.42 | -3.58 | -9.63 | No |
|  | Sex+Race | 0.6642 | 0.7645 | 1.58 | -4.42 | -13.49 | No |

FDR-adjusted $p < 0.05$ highlighted in red

**Supplementary Table 8.** Term-removal model comparison for SSC fluorescence lifetime parameters.

| Outcome | Term dropped | LRT p | FDR-adjusted p | LR statistic | ΔAIC | ΔBIC | Keep |
| --- | --- | --- | --- | --- | --- | --- | --- |
| SSC $\tau_{m12}$ | Age | 0.0000 | 0.0001 | 18.90 | 16.90 | 13.88 | Yes |
|  | Location | 0.0000 | 0.0000 | 125.53 | 123.53 | 120.51 | Yes |
|  | Sex | 0.5364 | 0.8295 | 0.38 | -1.62 | -4.64 | No |
|  | Race | 0.1054 | 0.6054 | 4.50 | 0.50 | -5.55 | No |
|  | Sex+Race | 0.1892 | 0.7645 | 4.77 | -1.23 | -10.30 | No |
| SSC $\tau_1$ | Age | 0.0000 | 0.0001 | 18.43 | 16.43 | 13.41 | Yes |
|  | Location | 0.0000 | 0.0000 | 115.12 | 113.12 | 110.10 | Yes |
|  | Sex | 0.4690 | 0.8295 | 0.52 | -1.48 | -4.50 | No |
|  | Race | 0.2871 | 0.6054 | 2.50 | -1.50 | -7.55 | No |
|  | Sex+Race | 0.4082 | 0.7645 | 2.89 | -3.11 | -12.18 | No |
| SSC $\tau_2$ | Age | 0.0086 | 0.0200 | 6.91 | 4.91 | 1.89 | Yes |
|  | Location | 0.0000 | 0.0000 | 46.80 | 44.80 | 41.77 | Yes |
|  | Sex | 0.6759 | 0.8295 | 0.17 | -1.83 | -4.85 | No |
|  | Race | 0.2464 | 0.6054 | 2.80 | -1.20 | -7.25 | No |
|  | Sex+Race | 0.4074 | 0.7645 | 2.90 | -3.10 | -12.17 | No |
| SSC $\alpha_1$ | Age | 0.0586 | 0.0911 | 3.58 | 1.58 | -1.45 | No |
|  | Location | 0.0058 | 0.0074 | 7.61 | 5.61 | 2.59 | Yes |
|  | Sex | 0.5708 | 0.8295 | 0.32 | -1.68 | -4.70 | No |
|  | Race | 0.5631 | 0.6890 | 1.15 | -2.85 | -8.90 | No |
|  | Sex+Race | 0.7099 | 0.7645 | 1.38 | -4.62 | -13.69 | No |
| SSC $\alpha_2$ | Age | 0.0455 | 0.0833 | 4.00 | 2.00 | -1.02 | No |
|  | Location | 0.0054 | 0.0074 | 7.74 | 5.74 | 2.71 | Yes |
|  | Sex | 0.6044 | 0.8295 | 0.27 | -1.73 | -4.76 | No |
|  | Race | 0.5236 | 0.6890 | 1.29 | -2.71 | -8.75 | No |
|  | Sex+Race | 0.6881 | 0.7645 | 1.47 | -4.53 | -13.60 | No |

FDR-adjusted p < 0.05 highlighted in red

**Supplementary Table 9.**
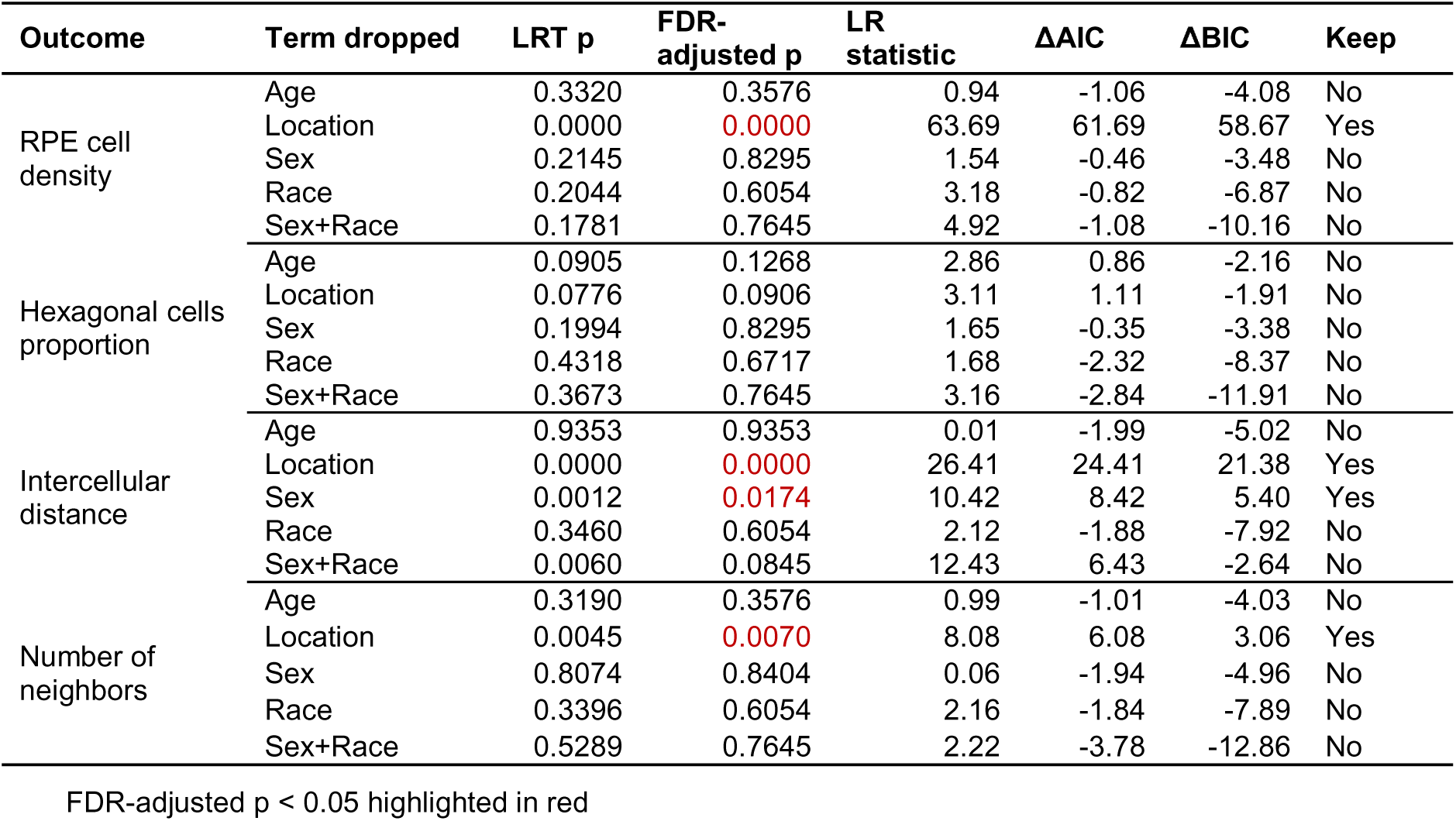
Term-removal model comparison for cellular structural metrics.

| Outcome | Term dropped | LRT p | FDR-adjusted p | LR statistic | ΔAIC | ΔBIC | Keep |
| --- | --- | --- | --- | --- | --- | --- | --- |
| RPE cell density | Age | 0.3320 | 0.3576 | 0.94 | -1.06 | -4.08 | No |
|  | Location | 0.0000 | 0.0000 | 63.69 | 61.69 | 58.67 | Yes |
|  | Sex | 0.2145 | 0.8295 | 1.54 | -0.46 | -3.48 | No |
|  | Race | 0.2044 | 0.6054 | 3.18 | -0.82 | -6.87 | No |
|  | Sex+Race | 0.1781 | 0.7645 | 4.92 | -1.08 | -10.16 | No |
| Hexagonal cells proportion | Age | 0.0905 | 0.1268 | 2.86 | 0.86 | -2.16 | No |
|  | Location | 0.0776 | 0.0906 | 3.11 | 1.11 | -1.91 | No |
|  | Sex | 0.1994 | 0.8295 | 1.65 | -0.35 | -3.38 | No |
|  | Race | 0.4318 | 0.6717 | 1.68 | -2.32 | -8.37 | No |
|  | Sex+Race | 0.3673 | 0.7645 | 3.16 | -2.84 | -11.91 | No |
| Intercellular distance | Age | 0.9353 | 0.9353 | 0.01 | -1.99 | -5.02 | No |
|  | Location | 0.0000 | 0.0000 | 26.41 | 24.41 | 21.38 | Yes |
|  | Sex | 0.0012 | 0.0174 | 10.42 | 8.42 | 5.40 | Yes |
|  | Race | 0.3460 | 0.6054 | 2.12 | -1.88 | -7.92 | No |
|  | Sex+Race | 0.0060 | 0.0845 | 12.43 | 6.43 | -2.64 | No |
| Number of neighbors | Age | 0.3190 | 0.3576 | 0.99 | -1.01 | -4.03 | No |
|  | Location | 0.0045 | 0.0070 | 8.08 | 6.08 | 3.06 | Yes |
|  | Sex | 0.8074 | 0.8404 | 0.06 | -1.94 | -4.96 | No |
|  | Race | 0.3396 | 0.6054 | 2.16 | -1.84 | -7.89 | No |
|  | Sex+Race | 0.5289 | 0.7645 | 2.22 | -3.78 | -12.86 | No |

FDR-adjusted p < 0.05 highlighted in red

### Supplementary note 7. Group-based mixed-effects analyses of AOFLIO outcomes in AMD

To quantify group- and lesion-specific differences in AOFLIO-derived outcomes in AMD eyes, linear mixed-effects models were fitted for each outcome with group as a fixed effect of interest. Pairwise contrasts were computed between healthy control regions (H), non-lesion AMD regions (NL), and distinct AMD lesion types, including soft drusen, subretinal drusenoid deposits (SDD), geographic atrophy (GA), acquired vitelliform lesions (AVL), and basal deposits (BD). All models included age and retinal eccentricity as covariates and a subject-specific random intercept to account for repeated measurements within individuals. Reported results include fixed-effect estimates (β), standard errors, test statistics, degrees of freedom, two-sided p values, and 95% confidence intervals for each contrast. Results are presented separately for LSC fluorescence lifetime parameters (**Supplementary Table 10**), SSC fluorescence lifetime parameters (**Supplementary Table 11**), and cellular structural metrics (**Supplementary Table 12**).

**Supplementary Table 10.** Group-based mixed-effects contrasts for LSC fluorescence lifetime parameters in AMD eyes.

| Outcome | Contrast | Estimate | SE | tStat | DF | pValue | CI- | CI+ |
| --- | --- | --- | --- | --- | --- | --- | --- | --- |
| LSC $\tau_{m12}$ | NL - H | 72.96 | 21.72 | 3.36 | 284 | 0.0009 | 30.20 | 115.71 |
|  | SDD - H | 81.81 | 22.48 | 3.64 | 284 | 0.0003 | 37.56 | 126.06 |
|  | SDD - NL | 8.85 | 9.38 | 0.94 | 284 | 0.3462 | -9.61 | 27.31 |
|  | GA - H | 467.31 | 47.73 | 9.79 | 284 | 0.0000 | 373.36 | 561.27 |
|  | GA - NL | 394.36 | 42.85 | 9.20 | 284 | 0.0000 | 310.00 | 478.71 |
|  | Drusen - H | 87.38 | 21.92 | 3.99 | 284 | 0.0001 | 44.22 | 130.54 |
|  | Drusen - NL | 14.42 | 8.02 | 1.80 | 284 | 0.0730 | -1.35 | 30.20 |
|  | AVL - H | 35.91 | 25.14 | 1.43 | 284 | 0.1542 | -13.56 | 85.39 |
|  | AVL - NL | -37.04 | 14.47 | -2.56 | 284 | 0.0110 | -65.53 | -8.56 |
|  | BD - H | 77.01 | 23.59 | 3.26 | 284 | 0.0012 | 30.57 | 123.45 |
|  | BD - NL | 4.05 | 11.37 | 0.36 | 284 | 0.7219 | -18.34 | 26.44 |
| LSC $\tau_1$ | NL - H | 34.60 | 9.76 | 3.55 | 284 | 0.0005 | 15.40 | 53.81 |
|  | SDD - H | 31.51 | 10.30 | 3.06 | 284 | 0.0024 | 11.23 | 51.79 |
|  | SDD - NL | -3.09 | 5.36 | -0.58 | 284 | 0.5642 | -13.64 | 7.45 |
|  | GA - H | 161.76 | 26.26 | 6.16 | 284 | 0.0000 | 110.08 | 213.45 |
|  | GA - NL | 127.16 | 24.59 | 5.17 | 284 | 0.0000 | 78.76 | 175.55 |
|  | Drusen - H | 34.70 | 9.89 | 3.51 | 284 | 0.0005 | 15.23 | 54.18 |
|  | Drusen - NL | 0.10 | 4.55 | 0.02 | 284 | 0.9828 | -8.86 | 9.06 |
|  | AVL - H | -7.94 | 12.14 | -0.65 | 284 | 0.5136 | -31.82 | 15.95 |
|  | AVL - NL | -42.54 | 8.26 | -5.15 | 284 | 0.0000 | -58.80 | -26.28 |
|  | BD - H | 35.17 | 11.06 | 3.18 | 284 | 0.0016 | 13.40 | 56.95 |
|  | BD - NL | 0.57 | 6.48 | 0.09 | 284 | 0.9300 | -12.19 | 13.33 |
| LSC $\tau_2$ | NL - H | 119.11 | 51.00 | 2.34 | 284 | 0.0202 | 18.74 | 219.49 |
|  | SDD - H | 157.44 | 53.51 | 2.94 | 284 | 0.0035 | 52.10 | 262.77 |
|  | SDD - NL | 38.32 | 26.26 | 1.46 | 284 | 0.1456 | -13.37 | 90.01 |
|  | GA - H | 843.37 | 129.75 | 6.50 | 284 | 0.0000 | 587.98 | 1098.76 |
|  | GA - NL | 724.26 | 120.31 | 6.02 | 284 | 0.0000 | 487.44 | 961.07 |
|  | Drusen - H | 124.89 | 51.65 | 2.42 | 284 | 0.0162 | 23.23 | 226.55 |
|  | Drusen - NL | 5.77 | 22.35 | 0.26 | 284 | 0.7964 | -38.23 | 49.77 |
|  | AVL - H | 131.11 | 62.07 | 2.11 | 284 | 0.0355 | 8.93 | 253.29 |
|  | AVL - NL | 11.99 | 40.49 | 0.30 | 284 | 0.7673 | -67.70 | 91.69 |
|  | BD - H | 84.79 | 57.08 | 1.49 | 284 | 0.1385 | -27.56 | 197.14 |
|  | BD - NL | -34.33 | 31.79 | -1.08 | 284 | 0.2811 | -96.89 | 28.24 |
| LSC $\alpha_1$ | NL - H | -3.06 | 1.41 | -2.18 | 284 | 0.0304 | -5.83 | -0.29 |
|  | SDD - H | -4.28 | 1.44 | -2.97 | 284 | 0.0032 | -7.12 | -1.44 |
|  | SDD - NL | -1.22 | 0.51 | -2.40 | 284 | 0.0169 | -2.21 | -0.22 |
|  | GA - H | -11.15 | 2.69 | -4.15 | 284 | 0.0000 | -16.44 | -5.86 |
|  | GA - NL | -8.09 | 2.31 | -3.50 | 284 | 0.0005 | -12.64 | -3.54 |
|  | Drusen - H | -4.70 | 1.42 | -3.32 | 284 | 0.0010 | -7.48 | -1.91 |
|  | Drusen - NL | -1.64 | 0.43 | -3.77 | 284 | 0.0002 | -2.49 | -0.78 |
|  | AVL - H | -3.36 | 1.56 | -2.15 | 284 | 0.0325 | -6.44 | -0.28 |
|  | AVL - NL | -0.30 | 0.78 | -0.38 | 284 | 0.7021 | -1.84 | 1.24 |
|  | BD - H | -4.70 | 1.49 | -3.15 | 284 | 0.0018 | -7.64 | -1.76 |
|  | BD - NL | -1.64 | 0.62 | -2.66 | 284 | 0.0082 | -2.85 | -0.43 |
| LSC $\alpha_2$ | NL - H | 2.63 | 1.27 | 2.06 | 284 | 0.0401 | 0.12 | 5.13 |
|  | SDD - H | 3.45 | 1.30 | 2.65 | 284 | 0.0086 | 0.88 | 6.02 |
|  | SDD - NL | 0.82 | 0.45 | 1.81 | 284 | 0.0707 | -0.07 | 1.72 |
|  | GA - H | 2.10 | 2.42 | 0.87 | 284 | 0.3855 | -2.66 | 6.86 |
|  | GA - NL | -0.53 | 2.07 | -0.25 | 284 | 0.7998 | -4.60 | 3.55 |
|  | Drusen - H | 3.95 | 1.28 | 3.08 | 284 | 0.0023 | 1.42 | 6.47 |
|  | Drusen - NL | 1.32 | 0.39 | 3.39 | 284 | 0.0008 | 0.55 | 2.09 |
|  | AVL - H | 1.97 | 1.41 | 1.39 | 284 | 0.1646 | -0.81 | 4.75 |
|  | AVL - NL | -0.66 | 0.70 | -0.94 | 284 | 0.3494 | -2.04 | 0.72 |
|  | BD - H | 3.77 | 1.35 | 2.79 | 284 | 0.0056 | 1.12 | 6.43 |
|  | BD - NL | 1.15 | 0.55 | 2.08 | 284 | 0.0386 | 0.06 | 2.23 |

P value less than 0.05 highlighted in red.

**Supplementary Table 11.**
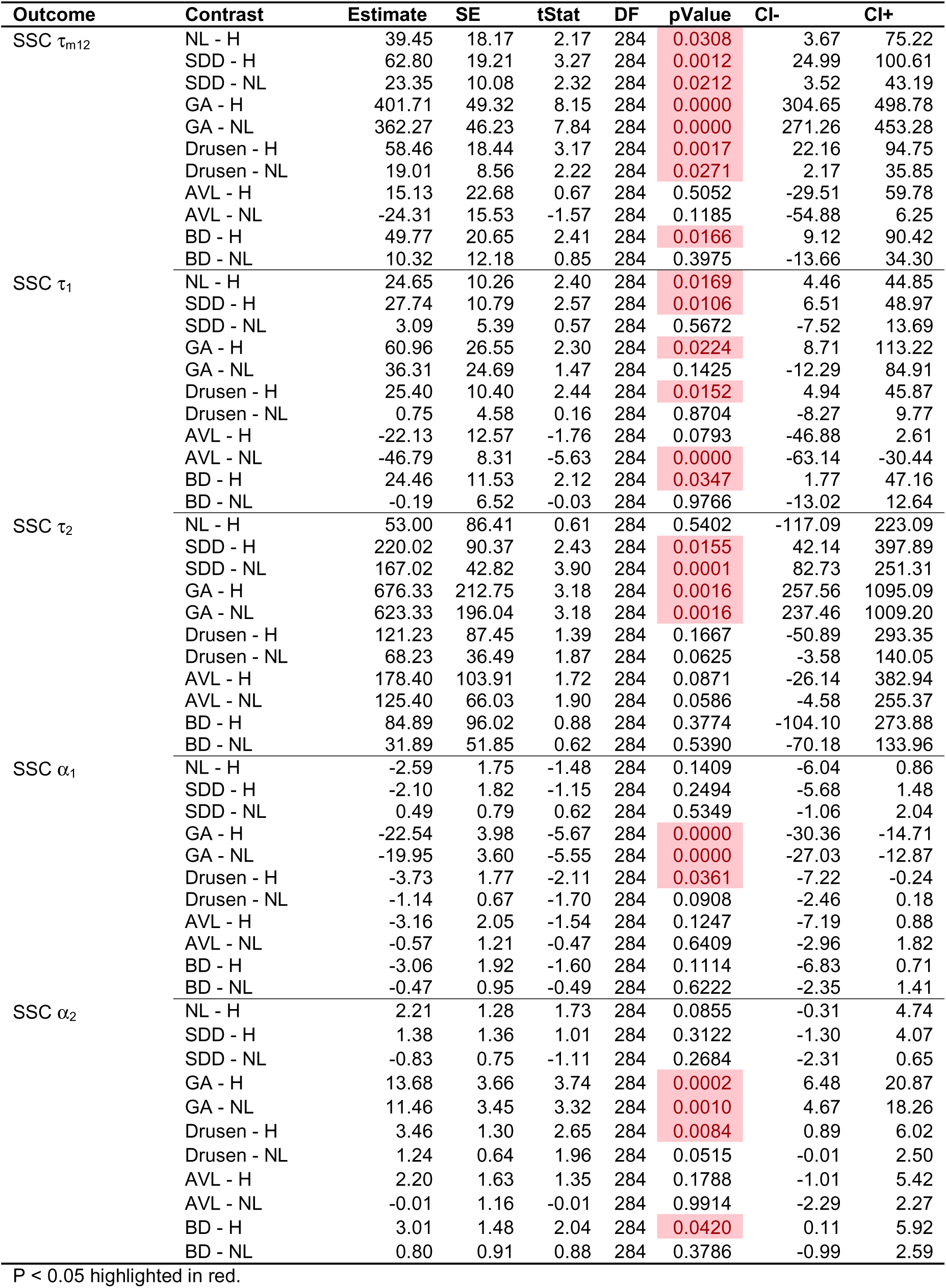
Group-based mixed-effects contrasts for SSC fluorescence lifetime parameters in AMD eyes.

**Supplementary Table 12.** Group-based mixed-effects contrasts for cellular structural metrics in AMD eyes.

| Outcome | Contrast | Estimate | SE | tStat | DF | pValue | CI- | CI+ |
| --- | --- | --- | --- | --- | --- | --- | --- | --- |
| RPE cell density | NL - H | -604.73 | 204.29 | -2.96 | 284 | 0.0033 | -1006.85 | -202.60 |
|  | SDD - H | -943.67 | 221.01 | -4.27 | 284 | 0.0000 | -1378.69 | -508.65 |
|  | SDD - NL | -338.94 | 136.58 | -2.48 | 284 | 0.0137 | -607.79 | -70.10 |
|  | GA - H | 452.50 | 657.77 | 0.69 | 284 | 0.4921 | -842.21 | 1747.22 |
|  | GA - NL | 1057.23 | 630.80 | 1.68 | 284 | 0.0948 | -184.40 | 2298.86 |
|  | Drusen - H | -638.92 | 208.25 | -3.07 | 284 | 0.0024 | -1048.83 | -229.00 |
|  | Drusen - NL | -34.19 | 115.01 | -0.30 | 284 | 0.7665 | -260.57 | 192.19 |
|  | AVL - H | -318.15 | 274.66 | -1.16 | 284 | 0.2477 | -858.78 | 222.48 |
|  | AVL - NL | 286.57 | 210.12 | 1.36 | 284 | 0.1737 | -127.01 | 700.16 |
|  | BD - H | -352.25 | 242.98 | -1.45 | 284 | 0.1483 | -830.53 | 126.03 |
|  | BD - NL | 252.48 | 164.48 | 1.54 | 284 | 0.1259 | -71.28 | 576.23 |
| Hexagonal cells proportion | NL - H | 0.00 | 0.02 | -0.19 | 284 | 0.8525 | -0.05 | 0.04 |
|  | SDD - H | 0.01 | 0.03 | 0.45 | 284 | 0.6552 | -0.04 | 0.07 |
|  | SDD - NL | 0.02 | 0.02 | 0.70 | 284 | 0.4835 | -0.03 | 0.07 |
|  | GA - H | -0.06 | 0.12 | -0.51 | 284 | 0.6097 | -0.31 | 0.18 |
|  | GA - NL | -0.06 | 0.12 | -0.48 | 284 | 0.6330 | -0.30 | 0.18 |
|  | Drusen - H | 0.01 | 0.02 | 0.31 | 284 | 0.7553 | -0.04 | 0.06 |
|  | Drusen - NL | 0.01 | 0.02 | 0.65 | 284 | 0.5185 | -0.02 | 0.05 |
|  | AVL - H | -0.06 | 0.04 | -1.42 | 284 | 0.1555 | -0.13 | 0.02 |
|  | AVL - NL | -0.05 | 0.04 | -1.45 | 284 | 0.1489 | -0.12 | 0.02 |
|  | BD - H | 0.05 | 0.03 | 1.47 | 284 | 0.1415 | -0.02 | 0.11 |
|  | BD - NL | 0.05 | 0.03 | 1.86 | 284 | 0.0636 | 0.00 | 0.11 |
| Intercellular distance | NL - H | 1.48 | 0.27 | 5.51 | 284 | 0.0000 | 0.95 | 2.01 |
|  | SDD - H | 2.12 | 0.29 | 7.33 | 284 | 0.0000 | 1.55 | 2.69 |
|  | SDD - NL | 0.64 | 0.18 | 3.66 | 284 | 0.0003 | 0.30 | 0.99 |
|  | GA - H | 0.48 | 0.85 | 0.57 | 284 | 0.5700 | -1.19 | 2.15 |
|  | GA - NL | -1.00 | 0.81 | -1.23 | 284 | 0.2194 | -2.60 | 0.60 |
|  | Drusen - H | 1.47 | 0.27 | 5.38 | 284 | 0.0000 | 0.93 | 2.01 |
|  | Drusen - NL | -0.01 | 0.15 | -0.06 | 284 | 0.9554 | -0.30 | 0.28 |
|  | AVL - H | 1.30 | 0.36 | 3.64 | 284 | 0.0003 | 0.60 | 2.01 |
|  | AVL - NL | -0.18 | 0.27 | -0.66 | 284 | 0.5119 | -0.71 | 0.35 |
|  | BD - H | 1.19 | 0.32 | 3.76 | 284 | 0.0002 | 0.57 | 1.82 |
|  | BD - NL | -0.29 | 0.21 | -1.35 | 284 | 0.1785 | -0.70 | 0.13 |
| Number of neighbors | NL - H | 0.05 | 0.02 | 2.22 | 284 | 0.0273 | 0.01 | 0.10 |
|  | SDD - H | 0.00 | 0.03 | 0.14 | 284 | 0.8886 | -0.05 | 0.06 |
|  | SDD - NL | -0.05 | 0.02 | -2.05 | 284 | 0.0417 | -0.09 | 0.00 |
|  | GA - H | -0.06 | 0.12 | -0.48 | 284 | 0.6330 | -0.29 | 0.18 |
|  | GA - NL | -0.11 | 0.12 | -0.92 | 284 | 0.3565 | -0.34 | 0.12 |
|  | Drusen - H | 0.02 | 0.02 | 0.66 | 284 | 0.5130 | -0.03 | 0.06 |
|  | Drusen - NL | -0.04 | 0.02 | -2.00 | 284 | 0.0462 | -0.07 | 0.00 |
|  | AVL - H | 0.01 | 0.04 | 0.23 | 284 | 0.8173 | -0.07 | 0.08 |
|  | AVL - NL | -0.04 | 0.03 | -1.27 | 284 | 0.2054 | -0.11 | 0.02 |
|  | BD - H | 0.06 | 0.03 | 2.05 | 284 | 0.0409 | 0.00 | 0.12 |
|  | BD - NL | 0.01 | 0.03 | 0.45 | 284 | 0.6547 | -0.04 | 0.06 |

P value less than 0.05 highlighted in red.

## Declaration

This manuscript describes original work that has not been published or submitted elsewhere.

## Ethics

The study followed the tenets of the Declaration of Helsinki, complied with the Health Insurance Portability and Accountability Act of 1996, and was approved by the Institutional Review Boards at the University of California at Los Angeles. Written informed consent was obtained from all participants.

## Data Availability

All data produced in the present work are contained in the manuscript.

## References

1 Feeney, L. Lipofuscin and melanin of human retinal pigment epithelium. Fluorescence, enzyme cytochemical, and ultrastructural studies. Investigative Ophthalmology & Visual Science 17, 583–600 (1978).

2 Eldred, G. E. & Katz, M. L. Fluorophores of the human retinal pigment epithelium: Separation and spectral characterization. Exp Eye Res 47, 71–86, 10.1016/0014-4835(88)90025-5 (1988).

3 Delori, F. C. Spectrophotometer for noninvasive measurement of intrinsic fluorescence and reflectance of the ocular fundus. Appl. Opt. 33, 7439–7452, doi:10.1364/ao.33.007439 (1994).

4 Delori, F. C. et al. In vivo fluorescence of the ocular fundus exhibits retinal pigment epithelium lipofuscin characteristics. Invest Ophthalmol Vis Sci 36, 718–729 (1995).

5 Schweitzer, D., Kolb, A. & Hammer, M. Autofluorescence lifetime measurements in images of the human ocular fundus. Vol. 4432 EBO (SPIE, 2001).

6 Elner, S. G. et al. Retinal flavoprotein autofluorescence as a measure of retinal health. Trans Am Ophthalmol Soc 106, 215–224 (2008).

7 Dysli, C. et al. Fluorescence lifetime imaging ophthalmoscopy. Prog Retin Eye Res 60, 120–143, doi:10.1016/j.preteyeres.2017.06.005 (2017).

8 Keilhauer, C. N. & Delori, F. o. C. Near-Infrared Autofluorescence Imaging of the Fundus: Visualization of Ocular Melanin. Investigative Ophthalmology & Visual Science 47, 3556–3564, doi:10.1167/iovs.06-0122 (2006).

9 Liu, T., Jung, H., Liu, J., Droettboom, M. & Tam, J. Noninvasive near infrared autofluorescence imaging of retinal pigment epithelial cells in the human retina using adaptive optics. Biomed Opt Express 8, 4348–4360, doi:10.1364/BOE.8.004348 (2017).

10 Sauer, L. et al. Review of clinical approaches in fluorescence lifetime imaging ophthalmoscopy. J Biomed Opt 23, 1–20, doi:10.1117/1.JBO.23.9.091415 (2018).

11 Schweitzer, D. et al. Towards metabolic mapping of the human retina. Microsc Res Tech 70, 410–419, doi:10.1002/jemt.20427 (2007).

12 Lienhart, W.-D., Gudipati, V. & Macheroux, P. The human flavoproteome. Archives of biochemistry and biophysics 535, 150–162, doi:10.1016/j.abb.2013.02.015 (2013).

13 Sparrow, J. R. & Boulton, M. RPE lipofuscin and its role in retinal pathobiology. Exp Eye Res 80, 595–606, doi:10.1016/j.exer.2005.01.007 (2005).

14 Ach, T. et al. Quantitative autofluorescence and cell density maps of the human retinal pigment epithelium. Invest Ophthalmol Vis Sci 55, 4832–4841, doi:10.1167/iovs.14-14802 (2014).

15 Ach, T. et al. Lipofuscin redistribution and loss accompanied by cytoskeletal stress in retinal pigment epithelium of eyes with age-related macular degeneration. Invest Ophthalmol Vis Sci 56, 3242–3252, doi:10.1167/iovs.14-16274 (2015).

16 Pollreisz, A. et al. Visualizing melanosomes, lipofuscin, and melanolipofuscin in human retinal pigment epithelium using serial block face scanning electron microscopy. Exp Eye Res 166, 131–139, doi:10.1016/j.exer.2017.10.018 (2018).

17 Bermond, K. et al. Autofluorescent Granules of the Human Retinal Pigment Epithelium: Phenotypes, Intracellular Distribution, and Age-Related Topography. Invest Ophthalmol Vis Sci 61, 35, doi:10.1167/iovs.61.5.35 (2020).

18 Pollreisz, A. et al. Atlas of Human Retinal Pigment Epithelium Organelles Significant for Clinical Imaging. Invest Ophthalmol Vis Sci 61, 13, doi:10.1167/iovs.61.8.13 (2020).

19 Kaarniranta, K. et al. Mechanisms of mitochondrial dysfunction and their impact on age-related macular degeneration. Prog Retin Eye Res, doi:10.1016/j.preteyeres.2020.100858 (2020).

20 Kong, G. Y. X., Van Bergen, N. J., Trounce, I. A. & Crowston, J. G. Mitochondrial Dysfunction and Glaucoma. Journal of Glaucoma 18 (2009).

21 Lefevere, E. et al. Mitochondrial dysfunction underlying outer retinal diseases. Mitochondrion 36, 66–76, doi:10.1016/j.mito.2017.03.006 (2017).

22 Berezin, M. Y. & Achilefu, S. Fluorescence lifetime measurements and biological imaging. Chemical reviews 110, 2641–2684 (2010).

23 Becker, W. The bh TCSPC Handbook. 8th edn, (Becker & Hickl GmbH, 2019).

24 Bernstein, P. S. et al. in High Resolution Imaging in Microscopy and Ophthalmology: New Frontiers in Biomedical Optics (ed Bille JF) (Springer, 2019).

25 Sauer, L. et al. Patterns of Fundus Autofluorescence Lifetimes In Eyes of Individuals With Nonexudative Age-Related Macular Degeneration. Invest Ophthalmol Vis Sci 59, AMD65–AMD77, doi:10.1167/iovs.17-23764 (2018).

26 Sauer, L. et al. Monitoring foveal sparing in geographic atrophy with fluorescence lifetime imaging ophthalmoscopy - a novel approach. Acta Ophthalmol 96, 257–266, doi:10.1111/aos.13587 (2018).

27 Dysli, C., Fink, R., Wolf, S. & Zinkernagel, M. S. Fluorescence Lifetimes of Drusen in Age-Related Macular Degeneration. Invest Ophthalmol Vis Sci 58, 4856–4862, doi:10.1167/iovs.17-22184 (2017).

28 Schweitzer, D. et al. Fluorescence lifetime imaging ophthalmoscopy in type 2 diabetic patients who have no signs of diabetic retinopathy. Journal of Biomedical Optics 20, 061106 (2015).

29 Dysli, C., Berger, L., Wolf, S. & Zinkernagel, M. S. Fundus autofluorescence lifetimes and central serous chorioretinopathy. *Retina (Philadelphia*, Pa.) 37, 2151–2161, doi:10.1097/IAE.0000000000001452 (2017).

30 Dysli, C., Schuerch, K., Escher, P., Wolf, S. & Zinkernagel, M. S. Fundus Autofluorescence Lifetime Patterns in Retinitis Pigmentosa. Invest Ophthalmol Vis Sci 59, 1769–1778, doi:10.1167/iovs.17-23336 (2018).

31 Dysli, C., Wolf, S., Tran, H. V. & Zinkernagel, M. S. Autofluorescence Lifetimes in Patients With Choroideremia Identify Photoreceptors in Areas With Retinal Pigment Epithelium Atrophy. Invest Ophthalmol Vis Sci 57, 6714–6721, doi:10.1167/iovs.16-20392 (2016).

32 Dysli, C., Wolf, S., Hatz, K. & Zinkernagel, M. S. Fluorescence Lifetime Imaging in Stargardt Disease: Potential Marker for Disease Progression. Invest Ophthalmol Vis Sci 57, 832–841, doi:10.1167/iovs.15-18033 (2016).

33 Sauer, L., Gensure, R. H., Hammer, M. & Bernstein, P. S. Fluorescence Lifetime Imaging Ophthalmoscopy: A Novel Way to Assess Macular Telangiectasia Type 2. *Ophthalmology*. Retina 2, 587–598, doi:10.1016/j.oret.2017.10.008 (2018).

34 Gantner, M. L. et al. Serine and Lipid Metabolism in Macular Disease and Peripheral Neuropathy. N Engl J Med 381, 1422–1433, doi:10.1056/NEJMoa1815111 (2019).

35 Sadda, S. R. et al. A pilot study of fluorescence lifetime imaging ophthalmoscopy in preclinical Alzheimer’s disease. Eye 33, 1271–1279, doi:10.1038/s41433-019-0406-2 (2019).

36 Williams, D. R., Burns, S. A., Miller, D. T. & Roorda, A. Evolution of adaptive optics retinal imaging [Invited]. Biomed Opt Express 14, 1307–1338, doi:10.1364/BOE.485371 (2023).

37 Burns, S. A., Elsner, A. E., Sapoznik, K. A., Warner, R. L. & Gast, T. J. Adaptive optics imaging of the human retina. Prog Retin Eye Res 68, 1–30, doi:10.1016/j.preteyeres.2018.08.002 (2019).

38 Bowles Johnson, K. E., et al. Fluorescence Lifetime Imaging of Human Retinal Pigment Epithelium in Pentosan Polysulfate Toxicity Using Adaptive Optics Scanning Light Ophthalmoscopy. Invest Ophthalmol Vis Sci 65, 27, doi:10.1167/iovs.65.4.27 (2024).

39 Kunala, K. et al. Near Infrared Autofluorescence Lifetime Imaging of Human Retinal Pigment Epithelium Using Adaptive Optics Scanning Light Ophthalmoscopy. Invest Ophthalmol Vis Sci 65, 27, doi:10.1167/iovs.65.5.27 (2024).

40 Kunala, K., Tang, J. A. H., Parkins, K. & Hunter, J. J. Multispectral label-free in vivo cellular imaging of human retinal pigment epithelium using adaptive optics fluorescence lifetime ophthalmoscopy improves feasibility for low emission analysis and increases sensitivity for detecting changes with age and eccentricity. J Biomed Opt 29, S22707, doi:10.1117/1.JBO.29.S2.S22707 (2024).

41 Tang, J. A. H. et al. Adaptive optics fluorescence lifetime imaging ophthalmoscopy of in vivo human retinal pigment epithelium. Biomed Opt Express 13, doi:10.1364/boe.451628 (2022).

42 Young, R. W. & Bok, D. Participation of the retinal pigment epithelium in the rod outer segment renewal process. J Cell Biol 42, 392–403, doi:10.1083/jcb.42.2.392 (1969).

43 Bok, D. The retinal pigment epithelium: a versatile partner in vision. Journal of cell science. Supplement 17, 189–195, doi:10.1242/jcs.1993.supplement_17.27 (1993).

44 Beatty, S., Koh, H., Phil, M., Henson, D. & Boulton, M. The role of oxidative stress in the pathogenesis of age-related macular degeneration. Surv Ophthalmol 45, 115–134, doi:10.1016/s0039-6257(00)00140-5 (2000).

45 Lamb, T. D. & Pugh, E. N., Jr. Dark adaptation and the retinoid cycle of vision. Prog Retin Eye Res 23, 307–380, doi:10.1016/j.preteyeres.2004.03.001 (2004).

46 Strauss, O. The retinal pigment epithelium in visual function. Physiol Rev 85, 845–881, doi:10.1152/physrev.00021.2004 (2005).

47 Lakowicz, J. R., Szmacinski, H., Nowaczyk, K. & Johnson, M. L. Fluorescence lifetime imaging of free and protein-bound NADH. Proc Natl Acad Sci U S A 89, 1271–1275, doi:10.1073/pnas.89.4.1271 (1992).

48 Sparrow, J. R. et al. The bisretinoids of retinal pigment epithelium. Prog Retin Eye Res 31, 121–135, doi:10.1016/j.preteyeres.2011.12.001 (2012).

49 Delori, F. o. C., Goger, D. G. & Dorey, C. K. Age-Related Accumulation and Spatial Distribution of Lipofuscin in RPE of Normal Subjects. Investigative Ophthalmology & Visual Science 42, 1855–1866 (2001).

50 Zhang, Y., Wang, X., Clark, M. E., Curcio, C. A. & Owsley, C. Imaging of Age-Related Macular Degeneration by Adaptive Optics Scanning Laser Ophthalmoscopy in Eyes With Aged Lenses or Intraocular Lenses. Transl Vis Sci Technol 9, 41, doi:10.1167/tvst.9.8.41 (2020).

51 Yu, Y., Zhang, T., Meadway, A., Wang, X. & Zhang, Y. High-speed adaptive optics for imaging of the living human eye. Opt. Express 23, 23035–23052, doi:10.1364/OE.23.023035 (2015).

52 Meadway, A., Girkin, C. A. & Zhang, Y. A dual-modal retinal imaging system with adaptive optics. Opt. Express 21, 29792–29807, doi:10.1364/OE.21.029792 (2013).

53 Yu, Y. & Zhang, Y. Dual-thread parallel control strategy for ophthalmic adaptive optics. Chinese optics letters : COL 12, 121202, doi:10.3788/col201412.121202 (2014).

54 Zhang, Y., Poonja, S. & Roorda, A. AOSLO: from benchtop to clinic. Vol. 6306 OP (SPIE, 2006).

55 Liu, R., Wang, X., Hoshi, S. & Zhang, Y. Substrip-based registration and automatic montaging of adaptive optics retinal images. Biomed Opt Express 15, 1311–1330, doi:10.1364/BOE.514447 (2024).

56 Fleckenstein, M., Schmitz-Valckenberg, S. & Chakravarthy, U. Age-Related Macular Degeneration: A Review. Jama 331, 147–157, doi:10.1001/jama.2023.26074 (2024).

57 Sarks, S. H., Arnold, J. J., Killingsworth, M. C. & Sarks, J. P. Early drusen formation in the normal and aging eye and their relation to age related maculopathy: a clinicopathological study. Br J Ophthalmol 83, 358–368, doi:10.1136/bjo.83.3.358 (1999).

58 Hageman, G. S. et al. An integrated hypothesis that considers drusen as biomarkers of immune-mediated processes at the RPE-Bruch’s membrane interface in aging and age-related macular degeneration. Prog Retin Eye Res 20, 705–732, doi:Doi 10.1016/S1350-9462(01)00010-6 (2001).

59 Curcio, C. A. et al. Subretinal drusenoid deposits in non-neovascular age-related macular degeneration: morphology, prevalence, topography, and biogenesis model. Retina 33, 265–276, doi:10.1097/IAE.0b013e31827e25e0 (2013).

60 Zweifel, S. A., Spaide, R. F., Curcio, C. A., Malek, G. & Imamura, Y. Reticular pseudodrusen are subretinal drusenoid deposits. Ophthalmology 117, 303–312.e301, doi:10.1016/j.ophtha.2009.07.014 (2010).

61 Wu, Z., Fletcher, E. L., Kumar, H., Greferath, U. & Guymer, R. H. Reticular pseudodrusen: A critical phenotype in age-related macular degeneration. Prog Retin Eye Res 88, 101017, doi:10.1016/j.preteyeres.2021.101017 (2022).

62 Chen, L., Messinger, J. D., Kar, D., Duncan, J. L. & Curcio, C. A. Biometrics, Impact, and Significance of Basal Linear Deposit and Subretinal Drusenoid Deposit in Age-Related Macular Degeneration. Invest Ophthalmol Vis Sci 62, 33, doi:10.1167/iovs.62.1.33 (2021).

63 Spaide, R. F. & Curcio, C. A. Drusen characterization with multimodal imaging. Retina 30, 1441–1454, doi:10.1097/IAE.0b013e3181ee5ce8 (2010).

64 Zhang, Y. et al. Photoreceptor perturbation around subretinal drusenoid deposits as revealed by adaptive optics scanning laser ophthalmoscopy. Am J Ophthalmol 158, 584–596 e581, doi:10.1016/j.ajo.2014.05.038 (2014).

65 Curcio, C. A. Soft Drusen in Age-Related Macular Degeneration: Biology and Targeting Via the Oil Spill Strategies. Invest Ophthalmol Vis Sci 59, AMD160-AMD181, doi:10.1167/iovs.18-24882 (2018).

66 Lindenberg, S. et al. Acquired Vitelliform Lesions in Intermediate Age-Related Macular Degeneration: A Cross Sectional Study. *Ophthalmology*. Retina 8, 854–862, doi:10.1016/j.oret.2024.04.009 (2024).

67 Sadda, S. R. et al. Consensus Definition for Atrophy Associated with Age-Related Macular Degeneration on OCT: Classification of Atrophy Report 3. Ophthalmology 125, 537–548, doi:10.1016/j.ophtha.2017.09.028 (2018).

68 Zhang, Y., Tiruveedhula, P., Sincich, L., Horton, J. & Roorda, A. Adaptive optics scanning laser ophthalmoscope (AOSLO) for precise visual stimulus presentation. J Vis 7, 116–116, doi:10.1167/7.15.116 (2007).

69 Sincich, L. C., Zhang, Y., Tiruveedhula, P., Horton, J. C. & Roorda, A. Resolving single cone inputs to visual receptive fields. Nat Neurosci 12, 967–969, doi:10.1038/nn.2352 (2009).

70 Harmening, W. M., Tiruveedhula, P., Roorda, A. & Sincich, L. C. Measurement and correction of transverse chromatic offsets for multi-wavelength retinal microscopy in the living eye. Biomed Opt Express 3, 2066–2077, doi:10.1364/boe.3.002066 (2012).

71 Winter, S. et al. Transverse chromatic aberration across the visual field of the human eye. J Vis 16, 9, doi:10.1167/16.14.9 (2016).

72 Yang, Q., Hunter, J. & Parkins, K. Microscopy imaging system and methods, U.S. Patent Application Publication US20230022632A1. U.S. Patent and Trademark Office. Microscopy imaging system and methods patent (January 26, 2023).

73 Curcio, C. A., Kar, D., Owsley, C., Sloan, K. R. & Ach, T. Age-Related Macular Degeneration, a Mathematically Tractable Disease. Invest Ophthalmol Vis Sci 65, 4, doi:10.1167/iovs.65.3.4 (2024).

74 Shirazi, M. F. et al. Visualizing human photoreceptor and retinal pigment epithelium cell mosaics in a single volume scan over an extended field of view with adaptive optics optical coherence tomography. Biomed Opt Express 11, 4520–4535, doi:10.1364/BOE.393906 (2020).

75 Kowalczuk, L. et al. in vivo Retinal Pigment Epithelium Imaging using Transscleral OPtical Imaging in healthy eyes. Ophthalmology Science, 100234 (2022).

76 Bower, A. J. et al. Integrating adaptive optics-SLO and OCT for multimodal visualization of the human retinal pigment epithelial mosaic. Biomed Opt Express 12, 1449–1466, doi:10.1364/BOE.413438 (2021).

77 Liu, Z. et al. Quantification of Human Photoreceptor-Retinal Pigment Epithelium Macular Topography with Adaptive Optics-Optical Coherence Tomography. Diagnostics (Basel) 14, doi:10.3390/diagnostics14141518 (2024).

78 Li, J. et al. Artificial intelligence assisted clinical fluorescence imaging achieves in vivo cellular resolution comparable to adaptive optics ophthalmoscopy. Commun Med (Lond) 5, 105, doi:10.1038/s43856-025-00803-z (2025).

79 Eppenberger, L. S. et al. Characterization of retinal pigment epithelium layer in healthy and diseased retinas with high-resolution adaptive optics transscleral flood illumination imaging. Acta Ophthalmol, doi:10.1111/aos.70016 (2025).

80 Ferdowsi, S. et al. In-Vivo Characterization of Healthy Retinal Pigment Epithelium and Photoreceptor Cells from AO-(T)FI Imaging. Vision 9, doi:10.3390/vision9040091 (2025).

81 Morgan, J. I., Dubra, A., Wolfe, R., Merigan, W. H. & Williams, D. R. In vivo autofluorescence imaging of the human and macaque retinal pigment epithelial cell mosaic. Invest Ophthalmol Vis Sci 50, 1350–1359, doi:10.1167/iovs.08-2618 (2009).

82 Grieve, K. et al. In vivo near-infrared autofluorescence imaging of retinal pigment epithelial cells with 757 nm excitation. Biomed Opt Express 9, 5946–5961, doi:10.1364/BOE.9.005946 (2018).

83 Granger, C. E. et al. Human Retinal Pigment Epithelium: In Vivo Cell Morphometry, Multispectral Autofluorescence, and Relationship to Cone Mosaic. Invest Ophthalmol Vis Sci 59, 5705–5716, doi:10.1167/iovs.18-24677 (2018).

84 Sparrow, J. R. & Duncker, T. Fundus Autofluorescence and RPE Lipofuscin in Age-Related Macular Degeneration. J Clin Med 3, 1302–1321, doi:10.3390/jcm3041302 (2014).

85 Borst, J. W., Hink, M. A., van Hoek, A. & Visser, A. J. Effects of refractive index and viscosity on fluorescence and anisotropy decays of enhanced cyan and yellow fluorescent proteins. J Fluoresc 15, 153–160, doi:10.1007/s10895-005-2523-5 (2005).

86 Lecinski, S. et al. Correlating viscosity and molecular crowding with fluorescent nanobeads and molecular probes: in vitro and in vivo. Interface Focus 12, 20220042, doi:10.1098/rsfs.2022.0042 (2022).

87 Islam, M. S., Honma, M., Nakabayashi, T., Kinjo, M. & Ohta, N. pH dependence of the fluorescence lifetime of FAD in solution and in cells. Int J Mol Sci 14, 1952–1963, doi:10.3390/ijms14011952 (2013).

88 Malacrida, L., Ranjit, S., Jameson, D. M. & Gratton, E. The Phasor Plot: A Universal Circle to Advance Fluorescence Lifetime Analysis and Interpretation. Annual Review of Biophysics 50, 575–593 (2021).

89 Smith, J. T. et al. Fast fit-free analysis of fluorescence lifetime imaging via deep learning. Proceedings of the National Academy of Sciences 116, 24019–24030 (2019).

90 Xiao, D., Sapermsap, N., Chen, Y. & Li, D. D. U. Deep learning enhanced fast fluorescence lifetime imaging with a few photons. Optica 10, 944–951 (2023).

91 Chen, A. X. et al. Functional imaging of mitochondria in retinal diseases using flavoprotein fluorescence. Eye (Lond) 35, 74–92, doi:10.1038/s41433-020-1110-y (2021).

92 Nimworaphan, J., Markowitz, D. M. & Sergott, R. C. Fluorescence lifetime imaging ophthalmoscopy adds the retina to cortical pathology for visual dysfunction in neurodegenerative diseases. Front Neurol 16, 1659264, doi:10.3389/fneur.2025.1659264 (2025).

93 Liu, R. et al. Evaluating the Influence of the Lens Autofluorescence on Adaptive Optics Fluorescence Lifetime Imaging Ophthalmoscopy. medRxiv : the preprint server for health sciences, doi:10.64898/2025.12.03.25341556 (2025).

94 Schweitzer, D., Haueisen, J., Brauer, J. L., Hammer, M. & Klemm, M. Comparison of algorithms to suppress artifacts from the natural lens in fluorescence lifetime imaging ophthalmoscopy (FLIO). Biomed Opt Express 11, 5586–5602, doi:10.1364/BOE.400059 (2020).

95 Lu, J., Gu, B., Wang, X. & Zhang, Y. High speed adaptive optics ophthalmoscopy with an anamorphic point spread function. Opt. Express 26, 14356–14374, doi:10.1364/OE.26.014356 (2018).

96 Staurenghi, G., Sadda, S., Chakravarthy, U. & Spaide, R. F. Proposed lexicon for anatomic landmarks in normal posterior segment spectral-domain optical coherence tomography: the IN•OCT consensus. Ophthalmology 10.1016/j.ophtha.2014.02.023 (2014).

